# Multi-centre derivation and validation of a colitis-associated colorectal cancer risk prediction web-tool

**DOI:** 10.1101/2020.04.10.20057869

**Authors:** Kit Curtius, Misha Kabir, Ibrahim Al Bakir, Chang-Ho Ryan Choi, Juanda L Hartono, Michael Johnson, James E East, Oxford IBD Cohort Study Investigators, James O Lindsay, Roser Vega, Siwan Thomas-Gibson, Janindra Warusavitarne, Ana Wilson, Trevor A Graham, Ailsa Hart

**Author notes:** joint first authors (Kit Curtius, Misha Kabir). joint senior authors (Ana Wilson, Trevor A Graham, Ailsa Hart). **Author contributions:** KC: study concept and design; acquisition of data; analysis and interpretation of data; drafting of the manuscript; critical revision of the manuscript for intellectual content; statistical analysis; software/web-tool development; obtained funding. MK: study concept and design; acquisition of data; analysis and interpretation of data; drafting of the manuscript; critical revision of the manuscript for intellectual content; statistical analysis. IAB: acquisition of data; analysis and interpretation of data. CHRC: acquisition of data; investigation of data. JH, MJ, Oxford IBD Cohort Study Investigators: acquisition of data. JEE: acquisition of data; interpretation of data; drafting of the manuscript; critical revision of the manuscript; obtained funding. JOL: acquisition of data; interpretation of data; drafting of the manuscript; critical revision of the manuscript. RV, JW: acquisition of data; interpretation of data; critical revision of the manuscript. STG: interpretation of data; drafting of the manuscript; critical revision of the manuscript. AW: study concept and design; study supervision; interpretation of data; drafting of the manuscript; critical revision of the manuscript for intellectual content. TAG: study concept and design; study supervision; obtained funding; interpretation of data; drafting of the manuscript; critical revision of the manuscript for intellectual content. AH: study concept and design; study supervision; interpretation of data; drafting of the manuscript; critical revision of the manuscript for intellectual content.

## Abstract

**Objective:** Ulcerative colitis (UC) patients diagnosed with low-grade dysplasia (LGD) have increased risk of developing advanced neoplasia (AN; high-grade dysplasia or colorectal cancer). We aimed to develop and validate a predictor of AN risk in UC patients with LGD and create a visual web-tool to effectively communicate the risk.

**Design:** In our retrospective multi-centre validated cohort study, adult UC patients with an index diagnosis of LGD, identified from four UK centres between 2001-2019, were followed until progression to AN. In the discovery cohort (n=246), a multivariate risk prediction model was derived from clinicopathological features using Cox regression. Validation used data from 3 external centres (n=198). The validated model was embedded in a web-tool to calculate patient-specific risk.

**Results:** Four clinicopathological variables were significantly associated with AN progression in the discovery cohort: endoscopically visible LGD > 1 cm (HR = 2.7; 95% CI 1.2-5.9), unresectable or incomplete endoscopic resection (HR = 3.4; 95% CI 1.6-7.4), moderate/severe histological inflammation within 5 years of LGD diagnosis (HR = 3.1; 95% CI 1.5-6.7), and multifocality (HR = 2.9; 95% CI 1.3-6.2). In the validation cohort, this 4-variable model accurately predicted future AN cases with overall calibration Observed/Expected = 1 (95% CI 0.63-1.5), and achieved 100% specificity for the lowest risk group over 13 years of available follow-up.

**Conclusion:** Multi-cohort validation confirms that patients with large, unresected, multifocal LGD and recent moderate/severe inflammation are at highest risk of developing AN. Personalised risk prediction provided via the Ulcerative Colitis-Cancer Risk Estimator *(www.UC-CaRE.uk)* can support treatment decision-making.

**SUMMARY BOX:** *What is already known about this subject?:*

- The risk of ulcerative colitis-associated low-grade dysplasia (LGD) progression to more advanced neoplasia is currently not clearly defined. The literature consists of historical data from small heterogenous observational studies with limited follow-up or lack of information on endoscopic resection status.

*What are the new findings?:*

- We present the results from the largest multi-centre cohort study to evaluate LGD long-term prognosis based on clinicopathological factors that are reflective of modern surveillance techniques.
- Recent moderate or severe active inflammation or LGD that is large, not fully resectable or is multifocal remain independent predictors of advanced neoplasia progression, even when stratified to reflect the most modern era of high definition endoscopic imaging, chromoendoscopy and advanced polypectomy techniques.
- Colorectal cancer incidence after endoscopic resection of unifocal polypoid and non-polypoid dysplasia is 0.6 per 100 patient-years.
- Long-term incidence of advanced neoplasia is similar if LGD is invisible or if LGD is visible but not completely endoscopically resected.
- Using these data, we have designed and externally validated a cancer risk prediction tool for ulcerative colitis patients with LGD.

*How might it impact on clinical practice in the foreseeable future?:*

- The *UC-CaRE* (Ulcerative Colitis-Cancer Risk Estimator) tool can be used to calculate and communicate individualised numerical cancer risk estimates to colitis patients with LGD.
- It facilitates the risk stratification of the lowest-risk patients, who can be reassured by undergoing continued surveillance, and those at the highest-risk who may benefit from a prophylactic colectomy.
- This visual aid presents the calculated risk in a graphical and pictorial form to optimise risk comprehension and informed treatment choices.

## Introduction

Patients with Ulcerative Colitis (UC) have an increased lifetime risk of developing colorectal cancer (CRC) and of CRC-related death^1–3^. Consequently, UC patients are advised to engage in a colonoscopic surveillance programme 8-10 years after diagnosis to detect and resect any dysplasia before it progresses to adenocarcinoma^4–7^. Due to a high CRC risk, high-grade dysplasia (HGD) warrants preventive colectomy surgery or e*n bloc* endoscopic resection with intensive surveillance follow-up if unifocal^4–7^. The natural history of low-grade dysplasia (LGD) progression is less well defined with a wide range of rates reported for progression of LGD lesions to advanced neoplasia (HGD or CRC) due to the inclusion of historical data from small population studies with heterogenous terminology and limited follow-up^8^. The quality of endoscopic surveillance has also evolved over the past three decades with standardisation of surveillance technique, advances in imaging technology such as high definition white-light imaging and chromoendoscopy, and resection techniques such as endoscopic mucosal resection and submucosal dissection. These advances have been linked with higher rates of visible dysplasia detection, lower proportions of dysplasia categorised as “invisible” and lower AN progression rates^8–10^. A recent systematic review of studies from the videoendoscopic era has found wide variation in AN progression rates after endoscopic resection from 0 – 23% at 5 years for polypoid LGD and 0 – 22% at 2 years for non-polypoid LGD^8^. A large observational study evaluating the effect of endoscopic resection on long-term LGD prognosis is required to be able to better inform patients of their cancer risk if they continue surveillance rather than have a colectomy.

Patients can be reluctant to consider surgical management even when the risks of CRC are high due to concerns about the negative impact that complications, stoma or ileoanal pouch function may have on their quality of life, given that they are often in clinical remission at the time of dysplasia detection^11–13^. Shared clinician-patient decision-making is particularly important when the evidence and best management option is unclear and there are potentially harmful consequences associated with the choice that is eventually made. This is the case for management of LGD in UC: the risks and consequences of developing CRC despite surveillance must be balanced against having a life-changing surgical operation. Providing evidence-based and individualised numerical CRC risk estimates using visual decision aids can promote patient engagement with decision-making^13–15^.

We aimed to evaluate the impact of endoscopic resection of LGD on risk of future AN in the largest multi-centre cohort study of this to date, identify the factors that can predict AN progression in the 21^st^ century and create an online simple and visual multivariate risk prediction model to communicate patient-specific risk. We created the Ulcerative Colitis-Cancer Risk Estimator (*UC-CaRE*) web-based application that is publicly accessible at www.uc-care.uk and can be used by clinicians to aid neoplasia risk communication, patient education and shared decision-making.

## Materials and Methods

### Study design and patient cohort identification

A retrospective cohort study of Ulcerative Colitis (UC) patients diagnosed with an index case of low-grade dysplasia (LGD) at four UK hospitals was undertaken. Hospital pathology databases were searched using the following terms to identify patients with UC who had been diagnosed with LGD: ‘ulcerative colitis’ or ‘inflammatory bowel disease’ and ‘dysplasia’, ‘low-grade dysplasia’, ‘adenocarcinoma’ or ‘dysplasia associated mass lesion (DALM)’. The searched time periods were marginally different between each site and started between 1^st^ January 2001 to 1^st^ January 2004 and ended between 31^st^ December 2016 to 31^st^ March 2019 (Figure 1).

**Figure 1:**
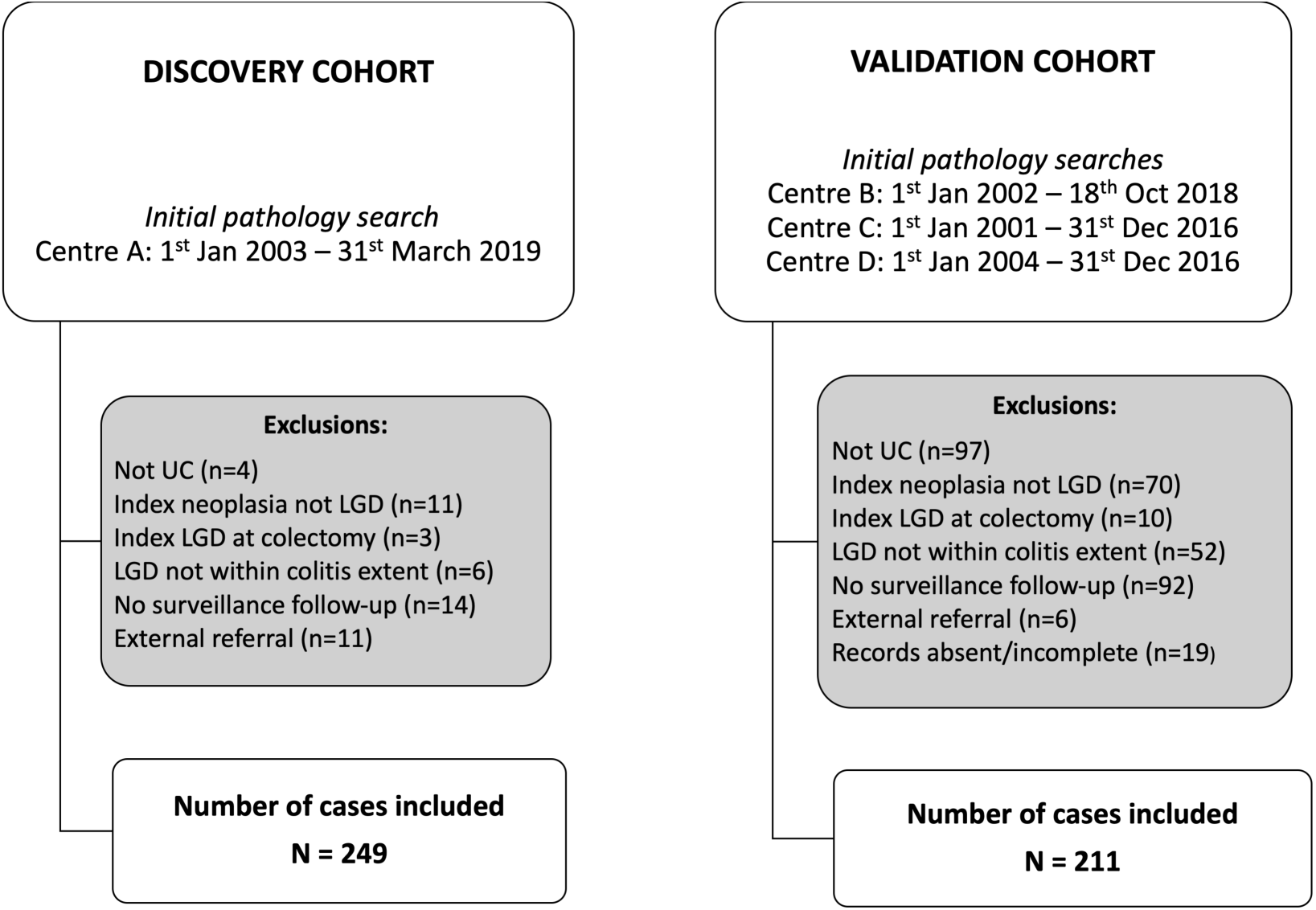
Flowchart of low-grade dysplasia (LGD) cases included and excluded (centre details in Table S1).

#### Inclusion criteria

Adult patients (aged 18 years or over) with histologically confirmed UC who had an index LGD diagnosis and had at least one follow-up examination of the whole colon after the index LGD diagnosis, either by colonoscopy or pathological analysis of a surgical colectomy specimen. The index LGD diagnosis was interpreted as the first documentation of LGD to have: 1) been confirmed by a second gastrointestinal histopathologist; 2) developed within the known histological extent of colitis based on historical pathology reports and 3) resection status known by judgement of the histopathologist/endoscopist.

#### Exclusion criteria

Patients were excluded if they had a diagnosis of Crohn’s disease, IBD-unclassified or indeterminate colitis, there was no adequate follow-up examination of the colon after the index LGD diagnosis, and the index LGD was: 1) located proximal to the known extent of historical microscopic inflammation as these were classed as sporadic adenomas; 2) diagnosed after a proctocolectomy i.e. was first noted incidentally within the surgical colonic specimen; 3) diagnosed at the same time as or after another more advanced neoplastic lesion (either high-grade dysplasia or adenocarcinoma); 4) diagnosed at an external institution to one of the study centres and the exact date of onset was unclear.

### Data collection

The clinical notes, endoscopy and histology reporting systems at each centre were interrogated to collect data for the following variables: patient age and duration of UC at time of index LGD diagnosis; patient gender; concomitant primary sclerosing cholangitis (which had been radiologically or histologically confirmed); patient exposure to 5-aminosalicylate, immunomodulators (thiopurines or methotrexate) and biological medications; macroscopic morphology of the index LGD as per the Paris classification^16^ (polypoid, non-polypoid or invisible); size and location of the largest visible index LGD; multifocality; completion of any endoscopic resection undertaken; presence of any macroscopic or histological active inflammation in the colon at the time of or within the 5 years preceding the index LGD; any chronic features of inflammation (colonic stricture, post-inflammatory polyps, scarred colon or a tubular and shortened colon); a previous diagnosis of indefinite for dysplasia; and use of chromoendoscopy during any surveillance colonoscopy performed before, at time of or after the index LGD diagnosis. Histological inflammation was categorised as moderate-severe active inflammation as defined by grade 3 or 4 of the Nancy histological index^17^. This requires the presence of ulceration and/or moderate to severe acute inflammatory cells infiltrate (multiple clusters of neutrophils in the lamina propria or epithelium). As reporting of the Nancy histological index or other similar validated histological inflammation scores was not standardised procedure across the centres in the study time period, a qualitative description correlating to Nancy histological index grade 3 or 4 was categorised as moderate-severe active inflammation. Cumulative inflammation burden (CIB), which has been shown to be an independent predictor of neoplasia incidence in IBD, was calculated as previously described^18^; namely as the sum of the average histological inflammation score between each pair of surveillance episodes multiplied by the surveillance interval in years^18^. A minimum of one documented colonic histological examination within 5 years prior to the index LGD diagnosis was required in order to be able to calculate a 5-year CIB.

Surveillance colonoscopy at each centre was performed in accordance with the national guidelines at the time^7,19^, including the use of chromoendoscopy. Dysplasia was categorised as invisible if it was detected on random mucosal biopsy with absence of a corresponding visible lesion in a colonic segment with good bowel preparation. If the lesion was found to be visible on targeted colonoscopy re-examination within 3 months, the lesion categorisation was changed from invisible to visible polypoid or non-polypoid. When multiple LGD lesions were found, categorisation of the morphology was based on the lesion considered to have more carcinogenic potential (from an earlier St Mark’s cohort^20^) in descending order of non-polypoid, invisible and polypoid morphology. The index LGD was categorised as multifocal LGD if more than one LGD discrete visible lesion was detected on index colonoscopy, regardless of the colonic segment, or foci of invisible LGD were detected in more than one colonic segment. Endoscopic resection where possible was based on histological confirmation of complete endoscopic resection, but often this could not be confirmed due to piecemeal resection or diathermy artefact, so completion of resection was based on endoscopic criteria. Invisible LGD was categorised as not being endoscopically resected. If there was multifocal LGD, whereby a visible lesion was successfully endoscopically resected but there was another focus of invisible LGD, this LGD case was categorised as not being successfully endoscopically resected. A lesion was considered a post-inflammatory polyp (PIP) if only inflammatory and/or granulation tissue and no neoplastic tissue was detected histologically within the lesion. Patients were recorded as having multiple PIPs if the endoscopist reported on there being ‘a few’, ‘several’, ‘many’ or ‘multiple’ PIPs within the colon. Colonic scarring was based on the endoscopist’s documentation of its macroscopic appearance.

### Follow-up outcomes

End of surveillance follow-up was determined by the date of the first incidence of advanced neoplasia (either HGD or CRC) or censoring at the last surveillance colonoscopy or proctocolectomy date.

### Statistical analysis of patient cohorts

The St Mark’s cohort of patients was used as the discovery set, and the patient cohorts from the three other centres were pooled together to form a validation set. Differences between the clinical characteristics of the cohorts were assessed using Chi-squared tests for categorical variables and Mann-Whitney U tests for non-parametric continuous variables (significance required p < 0.002 Bonferroni multiple testing correction). Data analysis was performed using SPSS (IBM SPSS Statistics for Macintosh, Version 25.0. Armonk, NY). Incidence rates of advanced neoplasia with 95% confidence interval [CI] were determined using OpenEpi software^21^.

### Statistical model selection and validation

In the discovery set, 17 clinicopathological variables were tested for associated advanced neoplasia risk using univariate Cox proportional hazard (PH) models (significance required p < 0.003 Bonferroni multiple testing correction). Significantly associated variables were included in a multivariate Cox PH model, and individual patient risk scores were computed. Kaplan-Meier (KM) estimation and log-rank tests were used to compare survival between dichotomised risk groups in discovery and validation sets. Positive and negative predictive values (PPV/NPV respectively) were assessed from KM curves to evaluate predictive power in the validation set. Survival analysis was carried out using the *survival* and *survminer* packages for R version 3.6.1. Estimation of cumulative incidence functions from the competing risk scenario of advanced neoplasia (AN) progression and colectomy during follow-up was performed using R package *cmprsk*.

### UC-CaRE risk prediction model development using discovery data

The *UC-CaRE* web tool was created to make patient-specific advanced neoplasia risk prediction, using Shiny interface to R.^22^ The multivariate model above was embedded in a web-tool that takes patient-specific features as user input, produces a cumulative advanced neoplasia (AN) risk curve into future years of follow-up, and displays a Paling chart illustrating the patient’s individual risk (see Supplementary material for prognostic risk function derivation).

### Evaluation of *UC-CaRE* risk predictions in validation data

We evaluated the risk predictions produced by the *UC-CaRE* tool in the independent validation dataset by computing the observed versus expected cumulative number of progressors to advanced neoplasia (AN) in the 13 years of follow-up data available post-baseline index LGD. The methods for assessing predictive model calibration are based on cumulative AN progression-specific hazards and may be used to assess calibration both overall and in risk score subgroups of the validation data (see Supplementary material for detailed model calibration methods).

### Ethical Considerations

The study was approved by the UK Research Ethics Committee (REC reference: 17/EM/0289; IRAS project ID: 227613).

## Results

### Study patient clinical characteristics and outcomes

A total of 460 patients were followed for 2,207 patient-years. There were 249 patients from St Mark’s Hospital (discovery cohort) and 211 patients in the multi-centre validation cohort (Figure 1) and detailed clinical characteristics were collected on each patient (Table S1, Figure S1). In the discovery cohort 7% (n=18/247) of the index LGD was invisible. Ninety percent (n = 209/231) of the visible LGD was successfully resected endoscopically. After LGD diagnosis, patients had a median of 4 follow-up colonoscopies (IQR 2.0 – 7.0) and a median follow up period of 5.1 years (IQR 2.3 – 8.5). Twenty percent (n = 50/249) eventually had a colectomy performed due to neoplasia or symptomatic disease after the index LGD diagnosis. Over the follow-up period, 7.2% (n = 18/249) progressed to CRC and a further 4.8% (n = 12/249) progressed to HGD only. Together, a total of 30 patients progressed to advanced neoplasia (AN).

There was significant heterogeneity in the clinicopathological characteristics between the discovery and validation cohorts as detailed in Table 1. Despite this, there were no significant differences in the incidence rates of advanced neoplasia (AN) and CRC between the two cohorts. The incidence rates of AN per 100 patient-years for the discovery cohort (n = 249), validation cohort (n = 211) and the total cohort (n = 460) were 2.2 (95% CI 1.5 - 3.0), 3.1 (95% CI 2.0 – 4.5) and 2.5 (95% CI 1.9 - 3.2) respectively. The incidence rates of CRC per 100 patient-years for the discovery cohort, validation cohort and the total cohort were 1.2 (95% CI 0.8 - 2.0), 1.8 (95% CI 1.0 - 2.9) and 1.5 per 100 patient-years (95% CI 1.0 - 2.1) respectively. The total cohort incidence rate of AN per 100 patient-years was: 1.1 (95% CI 0.7 – 1.8) after successful endoscopic resection of unifocal visible LGD; 5.2 (95% CI 1.9 - 11.5) if unifocal visible LGD was not completely endoscopically resected; 7.0 (95% CI 3.7 - 12.1) if there was unifocal invisible LGD; 2.6 (95% CI 1.3 – 4.8) after successful endoscopic resection of multifocal LGD; and 19.3 (95% CI 10.5 – 32.8) if multifocal LGD was not endoscopically resected. For the total cohort, incidence rates of CRC per 100 patient-years were: 0.6 (95% CI 0.3 - 1.1) after endoscopic resection of unifocal visible LGD; 2.0 (95% CI 0.3 - 6.7) if unifocal visible LGD was not successfully endoscopically resected; 4.3 (95% CI 1.9 - 8.6) if there was unifocal invisible LGD; 1.7 (95% CI 0.7 – 3.5) after successful endoscopic resection of multifocal LGD; and 12.6 (95% CI 5.9 – 24.0) if multifocal LGD was not endoscopically resected.

**Table 1:** Clinical characteristics of LGD patients for the discovery and validation cohorts. LGD = low-grade dysplasia; CRC = colorectal cancer; PSC = Primary sclerosing cholangitis. CIB = Cumulative inflammation burden score was defined as sum of: average score between each pair of surveillance episodes multiplied by the surveillance interval in years. Statistical significance required p < 0.002 with Bonferroni multiple testing correction.

| Clinical characteristics of included LGD patients | Discovery cohort (N = 249) | Validation cohort (N = 211) | Difference between groups (p-value) |
| --- | --- | --- | --- |
| **Gender** | (n = 249) | (n = 211) | 0.276 |
| - Female | 85 (34.1%) | 62 (29.4%) |  |
| - Male | 164 (65.9%) | 149 (70.6%) |  |
| **Median age at index LGD diagnosis (years)** | 62.0 (IQR 53.5 – 69.0) | 58.0 (IQR 48.0 – 68.0) | 0.021 |
| **Median duration of UC at index LGD diagnosis (years)** | 22.0 (IQR 12.0 – 33.0) | 16.0 (IQR 6.0 – 27.0) | <0.001 |
| **Colitis extent proximal to splenic flexure** | 225/249 (90.4%) | 166/210 (79.0%) | 0.001 |
| **Presence of concomitant PSC** | 13/248 (5.2%) | 38/191 (19.9%) | <0.001 |
| **Exposure to 5-aminosalicylates** | (n = 238) | (n = 74) | 0.382 |
| - None documented | 28 (11.8%) | 9 (12.2%) |  |
| - 0 – 10 years | 53 (22.3%) | 11 (14.9%) |  |
| - > 10 years | 157 (66.0%) | 54 (73.0%) |  |
| **Exposure to immunomodulators** | (n = 237) | (n = 75) | 0.354 |
| - None documented | 171 (72.2%) | 48 (64.0%) |  |
| - 0 – 10 years | 43 (18.1%) | 19 (25.3%) |  |
| - > 10 years | 23 (9.7%) | 8 (10.7%) |  |
| **Exposure to biological therapy** | 9/236 (3.8%) | 1/76 (1.3%) | 0.282 |
| **Morphology of index LGD** | (n = 247) | (n = 205) | <0.001 |
| - Polypoid | 142 (57.5%) | 133 (64.9%) |  |
| - Non-polypoid | 87 (35.2%) | 42 (20.5%) |  |
| - Invisible | 18 (7.3%) | 30 (14.6%) |  |
| **Visible index LGD size 10mm or more** | 79/248 (31.9%) | 68/202 (33.7%) | 0.684 |
| **Location of index LGD** | (n = 243) | (n = 209) | 0.001 |
| - Distal to splenic flexure | 115 (47.3%) | 133 (63.6%) |  |
| - Proximal to splenic flexure | 128 (52.7%) | 76 (36.4%) |  |
| **Successful endoscopic resection of index LGD (judged by endoscopic criteria)** | 209/249 (83.9%) | 139/207 (67.1%) | <0.001 |
| **Multifocal LGD at index diagnosis** | 61/249 (24.5%) | 37/211 (17.5%) | 0.069 |
| **Previous indefinite for dysplasia** | 12/249 (4.8%) | 9/210 (4.3%) | 0.785 |
| **Presence of a colonic stricture** | 8/249 (3.2%) | 2/210 (1.0%) | 0.098 |
| **Scarring/tubular/shortened colon** | 137/249 (55.0%) | 29/208 (13.9%) | <0.001 |
| **Multiple post-inflammatory polyps** | 88/241 (36.5%) | 58/209 (27.8%) | 0.048 |
| **Cumulative inflammation burden (CIB) score from 5 years preceding index LGD diagnosis** | (n=177)<br>1.5 (IQR 0.0-3.0) |  |  |
| **Maximum severity of histological active inflammation in any colonic segment at the same time or within previous 5 years of index LGD diagnosis** | (n = 247) | (n = 207) | 0.006 |
| - Quiescent | 105 (42.5%) | 66 (31.9%) |  |
| - Mild | 80 (32.4%) | 59 (28.5%) |  |
| - Moderate | 47 (19.0%) | 55 (26.6%) |  |
| - Severe | 15 (6.1%) | 27 (13.0%) |  |
| **Maximum severity of histological active inflammation in any colonic segment within 5 years after index LGD diagnosis** | (n = 227) | (n=207) | **0.003** |
| - Quiescent | 117 (51.5%) | 75 (36.2%) |  |
| - Mild | 52 (22.9%) | 54 (26.1%) |  |
| - Moderate | 47 (20.7%) | 53 (25.6%) |  |
| - Severe | 11 (4.8%) | 25 (12.1%) |  |
| **Chromoendoscopy use** | 202/245 (82.4%) | 148/208 (71.2%) | 0.004 |
| **Median no. of colonoscopies performed in surveillance follow-up** | 4.0 (IQR 2.0 – 7.0) | 2.0 (IQR 1.0 – 4.0) | **<0.001** |
| **Median follow-up time after index LGD diagnosis (years)** | 5.1 (IQR 2.2 – 8.5) | 3.1 (IQR 1.3 – 5.8) | **<0.001** |
| **Metachronous LGD on follow-up** | (n = 249) | (n = 209) | **<0.001** |
| - None | 84 (33.7%) | 109 (52.2%) |  |
| - In same colonic segment | 38 (15.3%) | 31 (14.8%) |  |
| - In different segment | 127 (51.0%) | 69 (33.0%) |  |
| **No. progressed to advanced neoplasia** | 30/249 (12.0%) | 25/211 (11.8%) | 0.948 |
| **No. progressed to CRC** | 18/249 (7.2%) | 15/211 (7.1%) | 0.960 |
| **Colectomy surgery after LGD diagnosis** | 50/249 (20.1%) | 38/211 (18.0%) | **0.001** |

A sub-analysis of the index LGD cases that were considered unresectable or incompletely resected was performed to determine whether differences in AN progression between visible and invisible LGD was due to differences in the proportion proceeding to proctocolectomy. In the situation where the highest malignant potential lesion detected was unresectable or incompletely resected non-polypoid LGD, 55.8% (n = 24/43) had proceeded to a proctocolectomy at time of censoring. In the situation where the highest malignant potential lesion detected was invisible LGD, proportionately fewer (39.6%; n = 12/48) had proceeded to proctocolectomy at time of censoring [*χ*^2^(1, N = 91) = 9.1; p=0.003]. Median time to proctocolectomy from index LGD diagnosis did not significantly differ between the invisible LGD group [11.5 months (IQR 3.8 - 21.0)] compared to the unresected non-polypoid LGD group [11.0 months (IQR 5.0 - 17.5)].

### Predictors of progression of low-grade dysplasia to advanced neoplasia (AN)

Five variables were found to be significantly predictive of progression to AN on univariate analysis of the discovery set (Table 2). Due to subjective inconsistencies in endoscopic reporting of macroscopic inflammation, two variables pertaining to histological inflammation status were used – presence of moderate or severe active histological inflammation (Nancy histological index^17^ grade 3 or 4 or equivalent) at the time of or within the previous 5 years of the index LGD diagnosis or the cumulative inflammation burden score [(CIB) the average histological inflammation score between each pair of surveillance episodes multiplied by the surveillance interval in years^18^]. The former was first entered into a multivariate model with the 3 other variables (Table 3, three patients removed due to missing LGD size or inflammation data). All four variables remained significant predictors of AN progression: size of any visible index LGD being 1cm or greater [adjusted HR 2.7 (95% CI 1.2 – 5.9); p=0.014]; unresectable or incomplete resection of the index LGD [adjusted HR 3.4 (95% CI 1.6 – 7.4); p=0.002]; multifocal LGD at index diagnosis [adjusted HR 2.9 (95% CI 1.3 – 6.2); p=0.007]; and presence of moderate or severe active histological inflammation at the time of or within the previous 5 years of the index LGD diagnosis [adjusted HR 3.1 (95% CI 1.5 – 6.7); p=0.003]. Partially due to less patient data available on CIB (n=177), we found that a 4-variable multivariate model using CIB as the alternative inflammation score variable was slightly less significant overall (Table S2) and data for CIB was not available for other centres so we utilised the first model for downstream analyses.

**Table 2.** Univariate analysis for progression to advanced neoplasia (AN) within the discovery set. Risk factors for low-grade dysplasia (LGD) progression to high-grade dysplasia or colorectal cancer (UNIVARIATE Cox regression analysis – 30/249 AN). Statistical significance required p < 0.003 with Bonferroni multiple testing correction. *Hazard ratio (HR) per 2-unit increase in cumulative inflammatory burden (equivalent to increase of 2 years continuous mild, 1 year continuous moderate or 8 months continuous severe active disease).

| Variable | Number (n=249) | Hazard ratio (95% CI) | P value |
| --- | --- | --- | --- |
| **Female sex** | 85 / 249 | 1.1 (0.5 – 2.3) | 0.790 |
| **Age at LGD diagnosis (years)** | (n = 249) |  |  |
| - Less than 40 | 16 | 1 |  |
| - 40 – 59 | 84 | 0.7 (0.2 – 3.4) | 0.708 |
| - 60 or more | 149 | 0.7 (0.2 – 3.1) | 0.656 |
| **Duration of UC at LGD diagnosis (years)** | (n = 247) |  |  |
| - 0 – 10 | 44 | 1 |  |
| - 11 – 20 | 70 | 3.4 (0.7 – 15.1) | 0.116 |
| - > 20 | 133 | 2.5 (0.6 – 10.6) | 0.229 |
| **Presence of concomitant PSC** | 13 / 248 | 2.6 (0.8 – 8.6) | 0.116 |
| **Patient exposure to 5-aminosalicylate medication** | (n = 238) |  |  |
| - None documented | 28 | 1 |  |
| - 0 – 10 years | 53 | 1.4 (0.3 – 5.4) | 0.664 |
| - > 10 years | 157 | 1.2 (0.4 – 4.2) | 0.729 |
| **Patient exposure to immunomodulator medication** | (n = 237) |  |  |
| - None documented | 171 | 1 |  |
| - 0 – 10 years | 43 | 2.4 (1.1 – 5.4) | 0.032 |
| - > 10 years | 23 | 0.8 (0.2 – 3.6) | 0.809 |
| **Macroscopic morphology of index LGD** | (n = 247) |  |  |
| - Polypoid | 142 | 1 |  |
| - Non-polypoid | 87 | 2.6 (1.2 – 5.7) | 0.016 |
| - Invisible | 18 | 4.1 (1.3 – 12.9) | 0.016 |
| **Visible index LGD size 10mm or more** | 79 / 248 | 3.8 (1.8 – 7.9) | **0.0004** |
| **Location of index LGD** | (n = 243) |  |  |
| - Distal to splenic flexure | 115 | 1 |  |
| - Proximal to splenic flexure | 128 | 0.7 (0.3 – 1.4) | 0.270 |
| **Index LGD not endoscopically resected or incomplete resection** | 40 / 249 | 4.7 (2.2 – 10) | **< 0.0001** |
| **Multifocal LGD at index diagnosis** | 61 / 249 | 3.2 (1.6 – 6.5) | **0.002** |
| **Previous diagnosis of indefinite for dysplasia** | 12 / 249 | 4.3 (1.5 – 12.5) | 0.007 |
| **Presence of a colonic stricture** | 8 / 249 | 5.5 (1.7 – 18.6) | 0.006 |
| **Scarring/tubular/shortened colon** | 137 / 249 | 1.6 (0.7 – 3.3) | 0.251 |
| **Moderate-severe histological active inflammation severity at time of or within previous 5 years of LGD diagnosis** | 62 / 247 | 3.6 (1.7 – 7.6) | **0.0006** |
| **Cumulative inflammation burden (CIB)\* (HR per 2 unit increase in CIB)** | (n=177) | 3.8 (1.8 – 8.0) | **0.0004** |
| **Multiple post-inflammatory polyps** | 88 / 241 | 1.5 (0.7 – 3.2) | 0.260 |

**Table 3.** Multivariate model for progression to advanced neoplasia (AN) within the discovery set. Risk factors for low-grade dysplasia (LGD) progression to high-grade dysplasia or colorectal cancer (MULTIVARIATE Cox regression analysis). N=246 total patients included with data available, 29 progressed to AN. Score (logrank) overall p=9e-10 for model.

| **Risk factor in final model** | **Hazard ratio (95% CI)** | **P value** |
| --- | --- | --- |
| **Visible index LGD size 10mm or more** | 2.7 (1.2 – 5.9) | **0.01400** |
| **Index LGD not endoscopically resected or incomplete resection** | 3.4 (1.6 – 7.4) | **0.00190** |
| **Multifocal LGD at time of index LGD diagnosis** | 2.9 (1.3 – 6.2) | **0.00749** |
| **Moderate or severe active histological inflammation in any colonic segment at time of or within previous 5 years of index LGD diagnosis** | 3.1 (1.5 – 6.7) | **0.00305** |

British Society of Gastroenterology (BSG) guidelines outlining dysplasia management were first published in 2010^19^. Since then there has also been increased adoption of chromoendoscopy, high definition imaging and advanced polypectomy techniques. To account for more recent changes in practice, we also performed a stratified multivariate analysis for index LGD diagnosed pre-2010 and in year 2010 or later. Fitted models assigned very similar risks to all four predictor variables in both eras (Table S3) and estimated similar baseline cumulative hazards estimated for each era (Figure S2). Thus, the adoption of modern endoscopic techniques does not appear to have altered features that define LGD progression risk.

To validate the multivariate model’s predictions, we turned to the validation set. Estimated baseline hazard was similar in both cohorts (Figure S5). Individual patient risk scores for the validation set were calculated and the predicted AN risk curves are illustrated in Figure S6. Risk prediction was similarly accurate in training and validation sets. We computed the observed (O) vs expected (E) standardized incidence ratio, O/E, on the validation set, finding O/E = 1 (95% CI 0.63 - 1.5) confirming the model’s efficacy and power in this independent data. Considering the combined modern data alone (LGD diagnosed in 2010 and later; 259 patients with all data on 4 risk factors), we found the model still accurately predicted the total number of 22 future AN cases, O/E = 1 (95% CI 0.63 - 1.5).

### Risk stratification with simple risk score in discovery and validation sets

We assigned a risk score to each patient based on the number of risk factors present (0 - 4 possible in total), combining patients with 3 or 4 risk factors due to low numbers. Kaplan-Meier (KM) curves for the risk tiers (Figure 2, Figure S3) in discovery vs validation sets confirmed very similar risk profiles in both cohorts (log-rank p < 0.0001 in both cohorts), which remained true when considering combined post-2010 data alone (Figure S4). Similar results were found when stratification was performed for 5 risk tiers (0-4) or 3 risk tiers (0, 1-2, 3+) (Figure S7).

**Figure 2:**
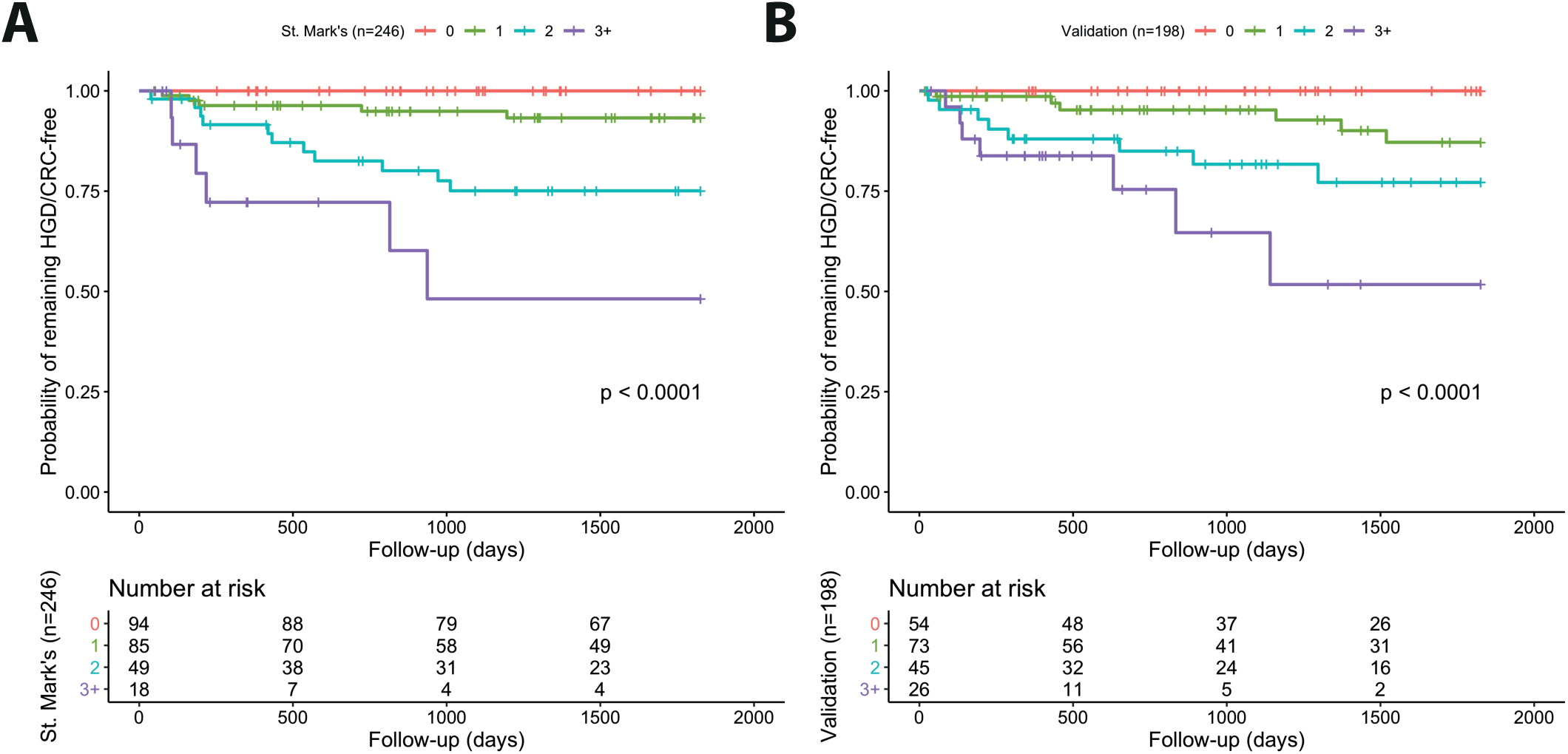
Kaplan-Meier plots for probability of remaining free of high-grade dysplasia (HGD) or colorectal cancer (CRC) to assess risk stratification and predictive power of multivariate model. (A) Discovery (n=246) and (B) validation (n=198) cohorts stratified by risk score (0-3+) defined by final multivariate model at index low-grade dysplasia (LGD) diagnosis to year 5 follow-up (see Figure S3 for similar results over total years of follow-up).

We computed predictive values for all risk groups in the discovery set and then found similar results for predictive power in the validation set (Table S4). Reassuringly, the group with lowest risk score = 0 (n = 54) in the validation set had a negative predictive value of 1 through all years of follow-up, i.e., no patient in this group progressed to AN, thus we determined lowest risk with perfect specificity using our model in this validation group. For the highest risk group, risk score = 3+ (n=26), in the validation set we found positive predictive values (PPV) of PPV = 12% by 6 months of follow-up, PPV = 16% by year 1, PPV = 35% by year 3, and PPV = 48% by year 5.

Finally, we conducted a competing risk analysis for time to colectomy versus risk of developing advanced neoplasia (AN). The hazard ratios, based on the four-risk factors above, were similar for both events (Figure S8). Thus, our findings suggest that our risk score predicts colectomy risk equivalently to predicting AN risk (Table S6 and Supplementary material)

### *UC-CaRE* risk prediction web-tool development

We built a web-tool named *UC-CaRE* to be used by a clinician to predict and display risk of AN for a UC patient with LGD. The tool takes the 4 patient-specific variables included in the final multivariate model as user input, and computes the function *Risk(t)* for probability of AN progression at time *t* based on those variables and the baseline hazard (see Materials and Methods and Supplementary material). Risk estimates are displayed as risk prediction curves (Figure 3), and also demonstrated with the aid of a diagram of 100 patients with the same risk, coloured according to how many of the total will likely develop an advanced neoplasm in 1, 5, and 10 years (Figure 4). This latter type of visual aid (also known as a Paling chart) can be helpful for patients to understand the meaning of a probability of cancer occurrence by viewing a simple diagram of predicted outcomes for 100 similarly at-risk UC patients (Figure 4). The ‘risk report’ summarizing the *UC-CaRE* output can be downloaded as a pdf file for ease of display and recording purposes.

**Figure 3:**
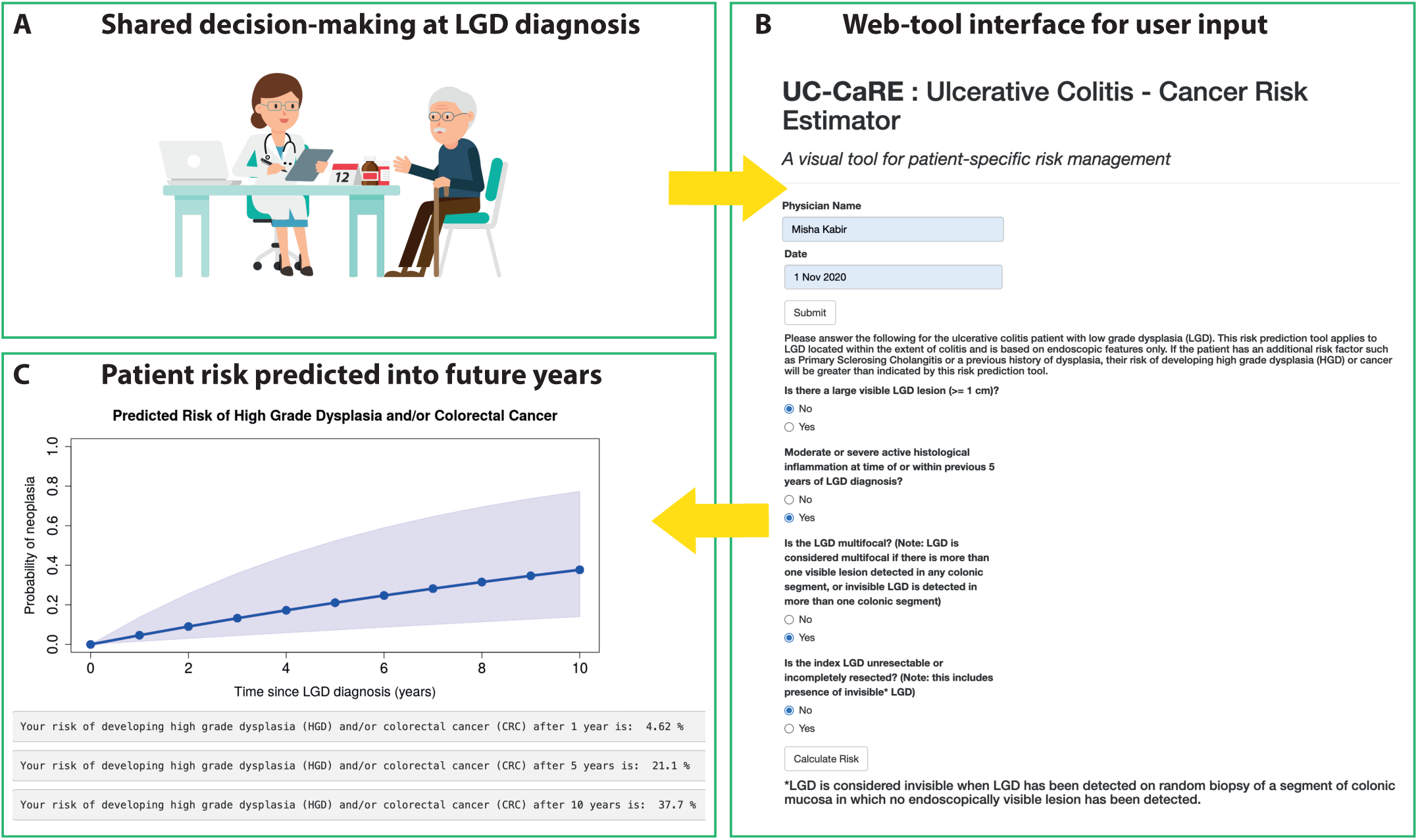
*UC-CaRE* clinical decision support web-tool user pipeline. A) Clinician records clinicopathological variables for patient at low-grade dysplasia (LGD) diagnosis (post-resection, if performed) for shared decision-making consultation. B) Simple interface in web-tool to input patient characteristics. C) Plot is created for predicted patient risk of progression to neoplasia at each year of future follow-up up to 10 years, with percentages also provided for consideration.

**Figure 4:**
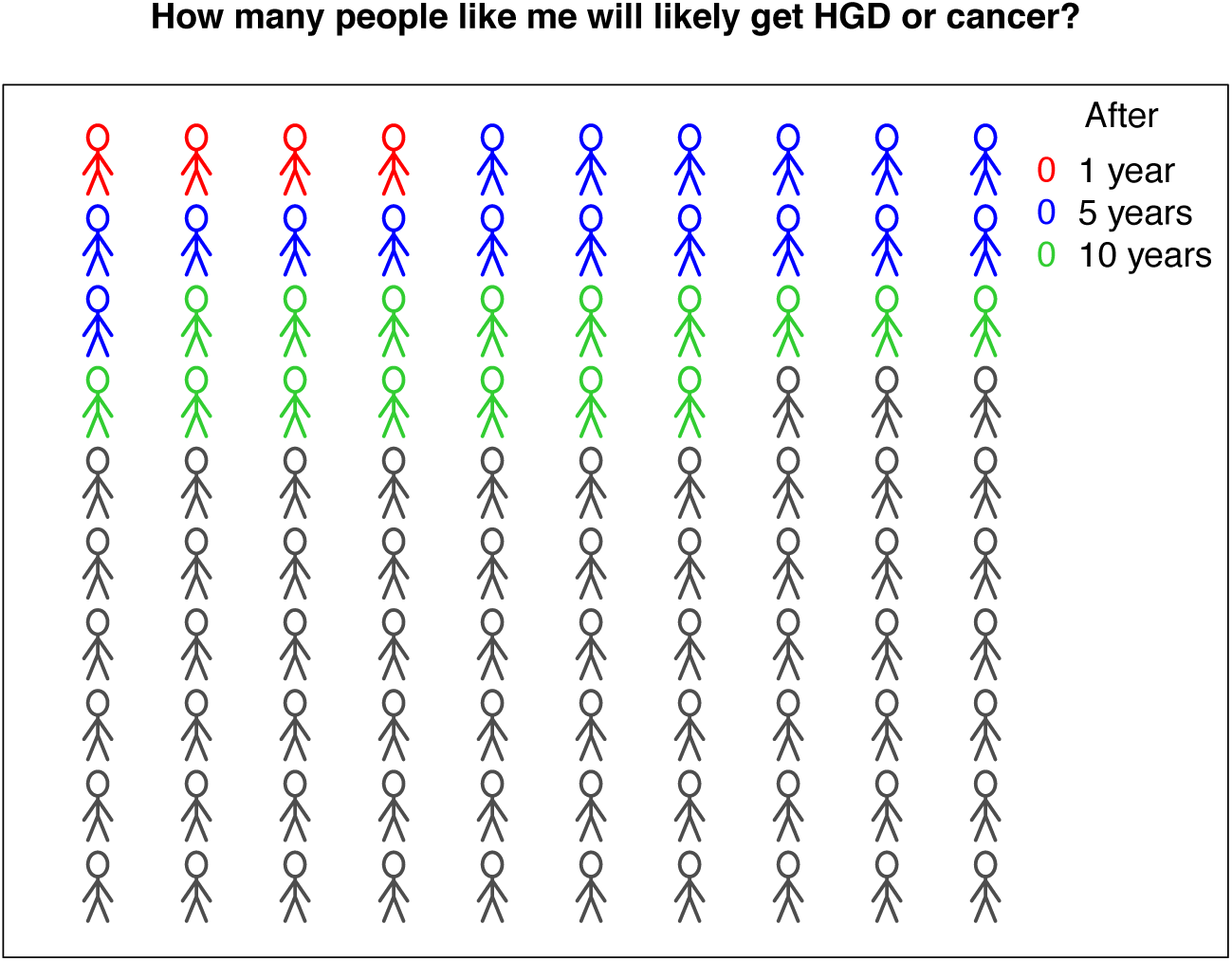
*UC-CaRE* online risk report. Paling charts provide a user-friendly display of the predicted cumulative risk of advanced neoplasia at 1, 5, and 10 years since low-grade dysplasia diagnosis/resection given patient characteristics.

## Discussion

We designed and validated a cancer risk prediction tool *UC-CaRE (*Ulcerative Colitis-Cancer Risk Estimator) using multi-centre data from UC patients diagnosed with low-grade dysplasia (LGD). We intend this tool to be used by clinicians to communicate personalised advanced neoplasia (AN) risk and to be used by patients to make a more informed choice to either accept colectomy or continue endoscopic surveillance.

Cancer risk communication in the context of LGD is particularly challenging as there remains much uncertainty about individualised risk. Communication of uncertainty or ambiguity in individualised CRC risk estimates can lead to an increase in patients avoiding decision-making^14^. Providing evidence-based and individualised numerical CRC risk estimates has been reported by patients to facilitate shared decision-making^13,15^. Visual decision aids that allow patients to view their individualised CRC risk in a graphical or pictorial form also promote patient risk comprehension and engagement with decision-making^14,23^. Clinicians currently lack an at-hand way to calculate and communicate an individual’s AN risk for dysplasia; *UC-CaRE* addresses this area of clinical need so that clinicians can make quantitative predictions of a UC patient’s risk of developing AN at point of care. The estimated absolute risk of AN at future years of surveillance can be easily demonstrated to a patient using the line graph and Paling chart automatically created by the web-tool.

We foresee a number of situations where the *UC-CaRE* tool could help decision-making in clinical practice. Patients can be reluctant to consider surgical management even when the risks of CRC are high due to concerns over a negative impact on their quality of life^11–13^. If a UC patient, for example was diagnosed with a 15mm non-polypoid endoscopically unresectable LGD lesion on a background of moderate active inflammation (i.e. has 3 risk factors), *UC-CaRE* predicts their 5 and 10-year risks would be 53% and 78% respectively and the high risk would appear visually apparent on the associated Paling chart. In the situation where one risk factor is modifiable i.e. the 15mm LGD lesion can be endoscopically resected or histological inflammatory quiescence is achieved, the 5 and 10-year CRC risks more than halve in the patient with 2 rather than 3 risk factors. In future scenarios where a new risk factor has appeared, i.e. multifocal LGD has developed, the UC-CaRE tool can be used to recalculate future CRC risk predictions. Importantly, the tool is able to predict with 100% specificity the patients at the lowest risk of progression to cancer and for whom continued surveillance rather than prophylactic colectomy can be confidently recommended.

The multivariate model we developed to embed in our tool was trained using current St Mark’s Hospital data, and then tested and validated in a dataset taken from three independent UK tertiary care centres specialising in inflammatory bowel disease. The model remained highly accurate in the validation set, even when only considering LGD diagnosed in the most recent post-2010 time period. We could predict progression for the first year of follow-up with positive predictive value 16% in the highest risk score group and negative predictive value 100% in the lowest risk score group; this information about risk will be very useful for patients faced with an imminent choice for management after diagnosis. We observed increasing accuracy at longer times in this cohort with positive predictive value reaching 48% by year 5 (and 35% by year 3), while perfect specificity of the lowest risk score group was sustained past 10 years of follow-up. This confirms that baseline findings are highly predictive in UC patients with LGD, and this should be considered when determining their personalised treatment and surveillance scheduling.

We recognised that censoring patients at colectomy, before they have had time to progress to advanced neoplasia (AN), was a competing risk for patients in our study. Our results confirm that the risk of both events (colectomy or AN progression) was similar, even when stratified by AN risk group. Thus, colectomy decisions made in the absence of an AN diagnosis are likely preventing AN development, with minimal over-treatment. Thus, it is reasonable to suggest that progression to AN could have been prevented by earlier colectomy in patients identified as high-risk by the *UC-CaRE* tool.

A number of clinicopathological variables have been reported by previous studies to be associated with AN progression after LGD diagnosis: patient specific characteristics such as age ≥ 55 years, male sex, follow-up at an academic (vs non-academic) medical centre, concomitant primary sclerosing cholangitis (PSC) and endoscopic characteristics of the index LGD such as non-polypoid morphology, invisibility (i.e. detected on random mucosal biopsy with no associated visible lesions), size greater than 10mm, multifocality, presence of a stricture, and distal location^20,24,25^. The St Mark’s patient dataset used in the discovery set overlaps with a previously reported cohort study of 172 UC patients with LGD diagnosed between 1993 to 2012 by Choi et al.^20^. In this older study, lesion size greater than 10mm (HR 10.0; 95% CI 4.3-23.4) and multifocality (HR 5.0; 95% CI 1.9-7.8) were significant predictive factors for AN progression. In Fumery et al.’s meta-analysis^25^ of LGD outcomes, multifocality (OR 3.5; 95% CI 1.5–8.5) was a significant predictive factor for AN development. In these two studies LGD morphology was found to be an additional risk factor on multivariate analysis. Choi et al.^20^ reported non-polypoid morphology (HR 16.5; 95% CI 6.8–39.8) and Fumery et al.^25^reported invisibility (OR 1.87; 95% CI 1.04–3.36) as being predictive of AN progression.

Strengths of our more recent study include the fact that it is the largest cohort study to date to have additionally evaluated the impact of endoscopic resectability on LGD prognosis. We note that endoscopic unresectability has not been assessed in previous studies,^20,24,25^ which is an important omission given that it is an indication for colectomy surgery to prevent CRC progression^4–7^. Another strength is the inclusion of LGD cases only diagnosed within the 21^st^ century, with additional analysis of the most recent post-2010 era. This is required to reflect recent advances in endoscopic surveillance. British Society of Gastroenterology (BSG) guidelines outlining IBD surveillance and dysplasia management were first published in 2010^19^. Chromoendoscopy was adopted into routine surveillance practice at all the study centres from the beginning of the study period and true high-definition imaging processors have been available from 2012 onwards. The development of advanced endoscopic resection techniques of non-polypoid dysplasia such as endoscopic submucosal dissection (ESD) and hybrid ESD/endoscopic mucosal resection (EMR), have allowed a greater number of these lesions to be endoscopically resected, when previously they would have been consigned to colectomy surgery^10^. More recent case series and smaller cohort studies from centres where high definition chromoendoscopy surveillance and advanced endoscopic resection techniques have been used have demonstrated lower rates of truly invisible dysplasia detection and lower AN progression rates after dysplasia has been endoscopically resected^8–10^. The univariate analyses in our study have indicated that whether or not a visible LGD lesion can be completely endoscopically resected is a more prominent risk factor than its morphology. We have demonstrated indistinguishable progression risk curves when comparing long-term AN incidence between truly invisible LGD and endoscopically unresectable or incompletely resected visible LGD in the validation data (Figure S9). We also found that a significantly greater proportion of the latter group proceeded to proctocolectomy before AN could develop. This practice reflects current guidelines which strongly advocate colectomy for unresectable visible dysplasia but acknowledge an uncertainty in the benefit of colectomy over continued surveillance for invisible dysplasia due to the low quality of evidence. Our study findings suggest that caution should be exercised if delaying proctocolectomy for invisible dysplasia.

We have reported incidence rates of LGD progression to advanced neoplasia (AN) and CRC of the total cohort (n=460) as being 2.5 and 1.5 per 100 patient-years follow-up respectively. These rates are higher than reported in Fumery et al.’s meta-analysis^25^ where the pooled AN and CRC incidence rates per 100-patients years were 1.8 and 0.8 respectively, but there was substantial calculated between-study heterogeneity (I^2^ statistic > 60%) and studies that included LGD proximal to the colitis extent were included. The inclusion of these latter cases may also explain why distal location of the LGD was found to be a predictive risk factor for progression to AN in the meta-analysis^25^ but not with our study cohort which only included LGD cases diagnosed within the extent of colitis. The incidence rate of CRC (1.4 per 100 patient-years) found in a Dutch population-based cohort study of 4284 IBD patients with LGD diagnosed at both academic and non-academic centres is very similar to ours^24^. There is a paucity of cohort studies reporting on long-term CRC incidence rates after endoscopic resection of non-polypoid dysplasia. Our incidence rate of CRC progression after endoscopic resection of both unifocal polypoid and non-polypoid dysplasia (0.6 per 100 patient-years) is very similar to the pooled incidence calculated in a meta-analysis of endoscopically resected polypoid-only dysplasia (0.5 per 100 patient-years)^26^.

Previous data from St Mark’s have demonstrated that the cumulative burden of inflammation (CIB) in the preceding years can predict the incidence of colonic neoplasia including large non-polypoid LGD, HGD and CRC [HR: 2.1 per 10-unit increase in CIB (equivalent of 10, 5 or 3.3 years of continuous mild, moderate or severe active histological inflammation); 95% CI 1.4 - 3.0; P<0.001]^18^. The predictive ability of the histologic CIB has been recently validated in an external centre^27^. This is in line with the theory that episodic inflammatory insults followed by mucosal healing encourage colonic epithelial cells to acquire carcinogenic genomic instability and mutations, conferring them a survival advantage to clonally proliferate and develop neoplastic lesions^28,29^. It is not surprising therefore that a significant proportion of the patients with LGD in this present study had experienced at least one episode of moderate-severe active histological inflammation in any colonic segment at the time of and within the preceding 5 years of the LGD diagnosis (25% in the discovery cohort and 40% in the validation cohort). Both this inflammation score variable and the 5-year CIB were predictive of LGD progression to advanced neoplasia (AN) progression. However, as several patients in the discovery cohort had not had a colonoscopy prior to the index LGD diagnosis, their CIB could not be calculated. Overall, the presence of an episode of moderate-severe active histological inflammation at the time of and within the preceding 5 years of the LGD diagnosis is a much more practical measure to compute for each patient than CIB and so we elected to use it in the final multivariate model rather than CIB.

It is important to note limitations of our study. This was a retrospective study relying on the accuracy of the available medical documentation, and incomplete medical records meant that other important risk factors for CRC development, such as smoking history and family history of CRC could not be included in the risk prediction model. Our study had only a modest number of PSC patients (n = 13) available to use in the discovery cohort. It is well recognised that PSC is an important risk for cancer progression in UC. Fumery et al.’s meta-analysis found that concomitant PSC (OR 3.4; 95% CI 1.5–7.8) was a significant risk factor for AN progression^25^, therefore it is likely *UC-CaRE* will underestimate AN risk in PSC patients and should not be used for this subset of patients. However, we note that increased AN risk in PSC was still predicted appropriately for the 38 PSC patients in the validation set because they had elevated risk predicted based on the same risk factors as the total UC population (for example, PSC patients had double the proportion of patients in the highest estimated risk group than that of the total UC population). By only including tertiary IBD centres in both discovery and validation cohorts, our results may also be limited by selection bias. However, we demonstrated significant heterogeneity between the two cohorts reflective of the variation in patient demographics and clinical practice likely to be seen outside of tertiary centres. Despite this heterogeneity we still found that the *UC-CaRE* model can accurately predict risk groups. We have also discussed above that the CRC incidence rates found with our study cohort were similar to that found in a population-based cohort study which included non-academic centres^24^. We therefore believe that the *UC-CaRE* model can be extrapolated to include patients form non-tertiary centres.

We also note significant proportions of the discovery and validation cohorts had experienced at least one episode of moderate-severe histological active inflammation in any colonic segment before, at the time of and/or even in the 5 years after the index LGD diagnosis. In addition, the only class of biological agent the patient cohorts in this study were exposed to was anti-TNF-alpha and overall exposure was low (3.8% in the discovery cohort and 1.3% in the validation cohort). This highlights how clinical practice and therapeutic targets have evolved over the time period of this study. The National Institute for Health and Care Excellence (NICE) only approved the use of anti-TNF-alpha agents in the UK for acute severe UC in 2008 and for chronic moderate-severe UC in 2015. Vedolizumab, ustekinumab and tofacitinib have since been approved for use in UC but none of our study patient cohorts were exposed to these. The benefits of achieving strict histological remission rather than relying on endoscopic or clinical symptom endpoints has also been realised in recent years with a shift in treatment targets^30^. We noted a trend for exposure to 5-aminosalicylates (less than 10 years duration) or immunomodulators to be associated with an increased risk of AN progression on univariate analyses. However, we believe that medication exposure in our study was a proxy for underlying chronic inflammation rather than AN progression occurring as a result of the medication exposure itself. Recent studies have suggested that immunomodulator^31^ and anti-TNF exposure^32^ may in fact be associated with lower cancer risk, but it is uncertain as to whether this is indirectly a result of achieving inflammatory control and mucosal healing or a direct chemoprotective effect. Further prospective data are required to understand the long-term impact of medical therapy, particularly biological drugs, on CRC risk in IBD.

## Conclusion

Our large multi-cohort retrospective validation study confirms that patients with large, unresected, and multifocal LGD and recent moderate/severe active inflammation are at the highest risk of developing advanced neoplasia. We have derived and validated a simple to use web-tool, *UC-CaRE (www.UC-CaRE.uk)*, for the calculation of personalised patient-specific high-grade dysplasia and/or CRC risk in individuals with UC and LGD. We believe the tool will be useful as an adjunct utilised by clinicians when managing CRC risk together with their patients. Further validation of the *UC-CaRE* tool using prospective data from non-tertiary centres will help test its applicability for generalised use and to reflect evolving clinical practice.

## Data Availability

All relevant clinical data summaries and statistical model outputs to reproduce results are supplied in the text, tables, and supplementary material.

## Acknowledgements

KC received funding from the MRC HDR-UK programme (UKRI Rutherford Fund Fellowship). TAG acknowledges funding from Cancer Research UK (A19771 and A25901) and the Barts Charity (472/2300). JEE was funded by the National Institute for Health Research (NIHR) Oxford Biomedical Research Centre. AL: Infrastructure support for this research was provided by the NIHR Imperial Biomedical Research Centre (BRC). The views expressed are those of the author(s) and not necessarily those of the National Health Service, the NIHR or the Department of Health. Oxford IBD cohort study investigators: Philip Allan, Tim Ambrose, Carolina Arancibia-Cárcamo, Adam Bailey, Ellie Barnes, Elizabeth Bird-lieberman, Jan Bornschein, Barbara Braden, Oliver Brain, Jane Collier, Emma Culver, James East, Alessandra Geremia, Bruce George, Lucy Howarth, Kelsey Jones, Paul Klenerman, Simon Leedham, Rebecca Palmer, Fiona Powrie, Astor Rodrigues, Jack Satsangi, Alison Simmons, Simon Travis, Holm Uhlig, Alissa Walsh

## Notes

**Conflict of interest statement:** KC, MK, JOL, JH, MJ, RV, STG, AW and TAG declare no potential conflicts of interest. JEE: Speaker FALK; consultant/shareholder Satisfai Health.

### Competing Interest Statement

KC, MK, JOL, JH, MJ, RV, STG, AW and TAG declare no potential conflicts of interest. JEE: Speaker FALK; consultant/shareholder Satisfai Health.

### Summary of Updates

Additional analyses added to enhance findings.

