## Supplementary material for "Multi-centre derivation and validation of a colitis-associated colorectal cancer risk prediction web-tool"

**Multi-centre derivation and validation of a colitis-associated colorectal cancer risk prediction web-tool:** *Supplementary methods, figures, and tables*

Authors: Kit Curtius[1*], Misha Kabir[2,3*], Ibrahim Al Bakir[1,2], Chang-Ho Ryan Choi[4], Juanda L Hartono[5,6], Michael Johnson[7,8], James E East[7,8], Oxford IBD Cohort Study Investigators[7,8], James O Lindsay[9,10], Roser Vega[11], Siwan Thomas-Gibson[2,3], Janindra Warusavitarne[2,3], Ana Wilson[2,3#], Trevor A Graham[1#], Ailsa Hart[2,3#]

Affiliations: [1]Centre for Genomics and Computational Biology, Barts Cancer Institute, Barts and the London School of Medicine and Dentistry, Queen Mary University of London, London, UK
[2]St Mark's Hospital, London, UK; [3]Imperial College London, UK
[4]St. George Hospital, Sydney, Australia
[5]Division of Gastroenterology, National University Hospital, Singapore
[6]Department of Medicine, Yong Loo Lin School of Medicine, National University of Singapore
[7]Translational Gastroenterology Unit, Nuffield Department of Medicine, John Radcliffe Hospital, University of Oxford, Oxford, UK
[8]Oxford NIHR Biomedical Research Centre, University of Oxford, Oxford, UK
[9]Centre for Immunobiology, Blizard Institute, Barts and the London School of Medicine and Dentistry, Queen Mary University of London, London, UK
[10]Barts Health NHS Trust, The Royal London Hospital, London, UK
[11]University College London Hospital, London, UK
* joint first authors
### joint senior authors

#### Supplementary Material

**Table S1. Clinical characteristics of LGD patients for each individual UK centre.**
JRH = John Radcliffe Hospital (centre B); UCLH = University College Hospital (centre C); RLH = Royal London Hospital (centre D); LGD = low-grade dysplasia; CRC = colorectal cancer; UC = Ulcerative colitis; PSC = Primary Sclerosing Cholangitis; IQR = interquartile range

| Clinical characteristics of included LGD patients | St Mark's (N = 249) | JRH (N = 135) | UCLH (N = 41) | RLH (N = 35) |
|---|---|---|---|---|
| **Gender** | (n = 249) | (n = 135) | (n = 41) | (n = 35) |
| - **Female** | 85 (34.1%) | 38 (28.1%) | 16 (39.0%) | 8 (22.9%) |
| - **Male** | 164 (65.9%) | 97 (71.9%) | 25 (61.0%) | 27 (77.1%) |
| **Median age at index LGD diagnosis (years)** | 62.0 (IQR 53.5 – 69.0) | 60.0 (IQR 49.0 – 69.0) | 62.0 (IQR 51.5 – 68.5) | 55.0 (IQR 44 – 60.0) |
| **Median duration of UC at index LGD diagnosis (years)** | 22.0 (IQR 12.0 – 33.0) | 16.0 (IQR 4.0 – 27.0) | 14.5 (IQR 5.8 – 27.0) | 18.0 (IQR 10 – 31) |
| **Colitis extends proximal to splenic flexure** | 225/249 (90.4%) | 104/134 (77.6%) | 34/41 (82.9%) | 28/35 (80.0%) |
| **Presence of concomitant PSC** | 13/248 (5.2%) | 28/119 (23.5%) | 7/40 (17.5%) | 3/32 (9.4%) |
| **Exposure to 5-aminosalicylates** | (n = 238) | Incomplete | (n = 39) | (n = 35) |
| - **None documented** | 28 (11.8%) | | 9 (23.1%) | 0 (0.0%) |
| - **0 – 10 years** | 53 (22.3%) | | 8 (20.5%) | 3 (8.6%) |
| - **> 10 years** | 157 (66.0%) | | 22 (56.4%) | 32 (91.4%) |
| **Exposure to immunomodulators** | (n = 237) | Incomplete | (n = 40) | (n = 35) |
| - **None documented** | 171 (72.2%) | | 27 (67.5%) | 21 (60.0%) |
| - **0 – 10 years** | 43 (18.1%) | | 10 (25.0%) | 9 (25.7%) |
| - **> 10 years** | 23 (9.7%) | | 3 (7.5%) | 5 (14.3%) |

| | | | | |
|---|---|---|---|---|
| **Exposure to biological therapy**<br>- **None documented**<br>- **0 – 10 years**<br>- **> 10 years** | (n = 236)<br>227 (96.2%)<br>9 (3.8%)<br>0 (0.0%) | Incomplete | (n = 41)<br>40 (97.6%)<br>1 (2.4%)<br>0 (0.0%) | (n = 35)<br>35 (100.0%)<br>0 (0.0%)<br>0 (0.0%) |
| **Morphology of index LGD**<br>- **Polypoid**<br>- **Non-polypoid**<br>- **Invisible** | (n = 247)<br>142 (57.5%)<br>87 (35.2%)<br>18 (7.3%) | (n = 133)<br>88 (66.2%)<br>25 (18.8%)<br>20 (15.0%) | (n = 37)<br>22 (59.5%)<br>11 (29.7%)<br>4 (10.8%) | (n = 35)<br>23 (65.7%)<br>6 (17.1%)<br>6 (17.1%) |
| **Visible index LGD size 10mm or more** | 79/248 (31.9%) | 41/111 (36.9%) | 17/32 (53.1%) | 10/29 (34.5%) |
| **Location of index LGD**<br>- **Distal to splenic flexure**<br>- **Proximal to splenic flexure** | (n = 243)<br>115 (47.3%)<br>128 (52.7%) | (n = 134)<br>88 (65.7%)<br>46 (34.3%) | (n = 41)<br>24 (58.5%)<br>17 (41.5%) | (n = 34)<br>21 (61.8%)<br>13 (38.2%) |
| **Successful endoscopic resection of index LGD (judged by endoscopic criteria)** | 209/249 (83.9%) | 85/132 (64.4%) | 29/40 (72.5%) | 25/35 (71.4%) |
| **Multifocal LGD at index diagnosis** | 61/249 (24.5%) | 24/135 (17.8%) | 9/41 (22.0%) | 4/35 (11.4%) |
| **Previous indefinite for dysplasia** | 12/249 (4.8%) | 3/135 (2.2%) | 3/40 (7.5%) | 3/35 (8.6%) |
| **Presence of a colonic stricture** | 8/249 (3.2%) | 0 (0.0%) | 2/40 (5.0%) | 0 (0.0%) |
| **Scarring/tubular/shortened colon** | 137/249 (55.0%) | 21/134 (15.7%) | 6/39 (15.4%) | 2/35 (5.7%) |
| **Multiple post-inflammatory polyps** | 88/241 (36.5%) | 32/134 (23.9%) | 12/40 (30.0%) | 14/35 (40.0%) |
| **Maximum severity of histological active inflammation in any colonic segment <u>at the same time or within previous 5 years</u> of index LGD diagnosis**<br>- **Quiescent**<br>- **Mild**<br>- **Moderate**<br>- **Severe** | (n = 247)<br>105 (42.5%)<br>80 (32.4%)<br>47 (19.0%)<br>15 (6.1%) | (n = 132)<br>46 (34.8%)<br>24 (18.2%)<br>39 (29.5%)<br>23 (17.4%) | (n = 41)<br>7 (17.1%)<br>19 (46.3%)<br>11 (26.8%)<br>4 (9.8%) | (n = 34)<br>13 (38.2%)<br>16 (47.1%)<br>5 (14.7%)<br>0 (0.0%) |
| **Maximum severity of histological active inflammation in any colonic segment <u>within 5 years after</u> index LGD diagnosis**<br>- **Quiescent**<br>- **Mild**<br>- **Moderate**<br>- **Severe** | (n = 227)<br>117 (51.5%)<br>52 (22.9%)<br>47 (20.7%)<br>11 (4.8%) | (n = 133)<br>53 (39.8%)<br>27 (53.0%)<br>31 (23.3%)<br>22 (16.5%) | (n = 40)<br>10 (25.0%)<br>13 (32.5%)<br>14 (35.0%)<br>3 (7.5%) | (n = 34)<br>12 (35.3%)<br>14 (41.2%)<br>8 (23.5%)<br>0 (0.0%) |
| **Cumulative inflammatory burden from 5 years preceding index LGD diagnosis** | 1.5<br>(IQR 0.0-3.0) | | | |
| **Chromoendoscopy use** | 202/245 (82.4%) | 92/135 (68.1%) | 29/38 (76.3%) | 27/35 (77.1%) |
| **Median follow-up time after index LGD diagnosis (years)** | 5.1<br>(IQR 2.2 – 8.5) | 3.2<br>(IQR 1.2 – 5.7) | 2.9<br>(IQR 1.4 – 5.6) | 2.8<br>(IQR 1.5 – 6.7) |
| **Metachronous LGD on follow-up**<br>- **None**<br>- **In same colonic segment**<br>- **In different segment** | (n = 249)<br>84 (33.7%)<br>38 (15.3%)<br>127 (51.0%) | (n = 134)<br>71 (53.0%)<br>20 (14.9%)<br>43 (32.1%) | (n = 41)<br>17 (41.5%)<br>8 (19.5%)<br>16 (39.0%) | (n = 34)<br>21 (61.8%)<br>3 (8.8%)<br>10 (29.4%) |
| **No. progressed to advanced neoplasia** | 30/249 (12.0%) | 15/125 (11.1%) | 7/41 (17.1%) | 3/35 (8.6%) |
| **No. progressed to CRC** | 18/249 (7.2%) | 8/135 (5.9%) | 5/41 (12.2%) | 2/35 (5.7%) |
| **Had colectomy surgery for dysplasia** | 50/249 (20.1%) | 31/135 (23.0%) | 13/41 (31.7%) | 6/35 (17.1%) |

**Table S2: Multivariate model for progression to advanced neoplasia (AN) within the discovery set using cumulative inflammation burden CIB) for inflammation score.**

Risk factors for low-grade dysplasia (LGD) progression to high-grade dysplasia or colorectal cancer (MULTIVARIATE Cox regression analysis). N=177 total patients considered with CIB, 24 progressed to AN. Score (logrank) overall p=2e-08 for model. *Adjusted hazard ratio (HR) per 2-unit increase in cumulative inflammatory burden (equivalent to increase of 2 years continuous mild, 1 year continuous moderate or 8 months continuous severe active disease).

| Risk factor in final model | Hazard ratio (95% CI) | P value |
|---|---|---|
| **Visible index LGD size 10mm or more** | 1.8 (0.8 – 4.3) | 0.16518 |
| **Index LGD not endoscopically resected or incomplete resection** | 3.6 (1.5 – 8.4) | 0.00373 |
| **Multifocal LGD at time of index LGD diagnosis** | 2.6 (1.1 – 6.1) | 0.02463 |
| **Cumulative inflammation burden (CIB)** | 1.5 (1.1 – 2.1) | 0.01079 |

**Figure S1: Violin plots comparing continuous clinical variables across patients in non-progressors ('no') vs progressors ('yes') to advanced neoplasia (HGD and/or CRC) outcome during study period in discovery set (n=249, top row) and validation set (n=211, bottom row).** Only significant differences were found in continuous size of largest low-grade dysplastic lesion in discovery set (NP median size = 5 mm vs. P median size = 15 mm, Mann-Whitney p = 0.0001) and in validation set (NP median size = 5 mm vs. P median size = 18 mm, p = 0.0027). LGD = low-grade dysplasia.

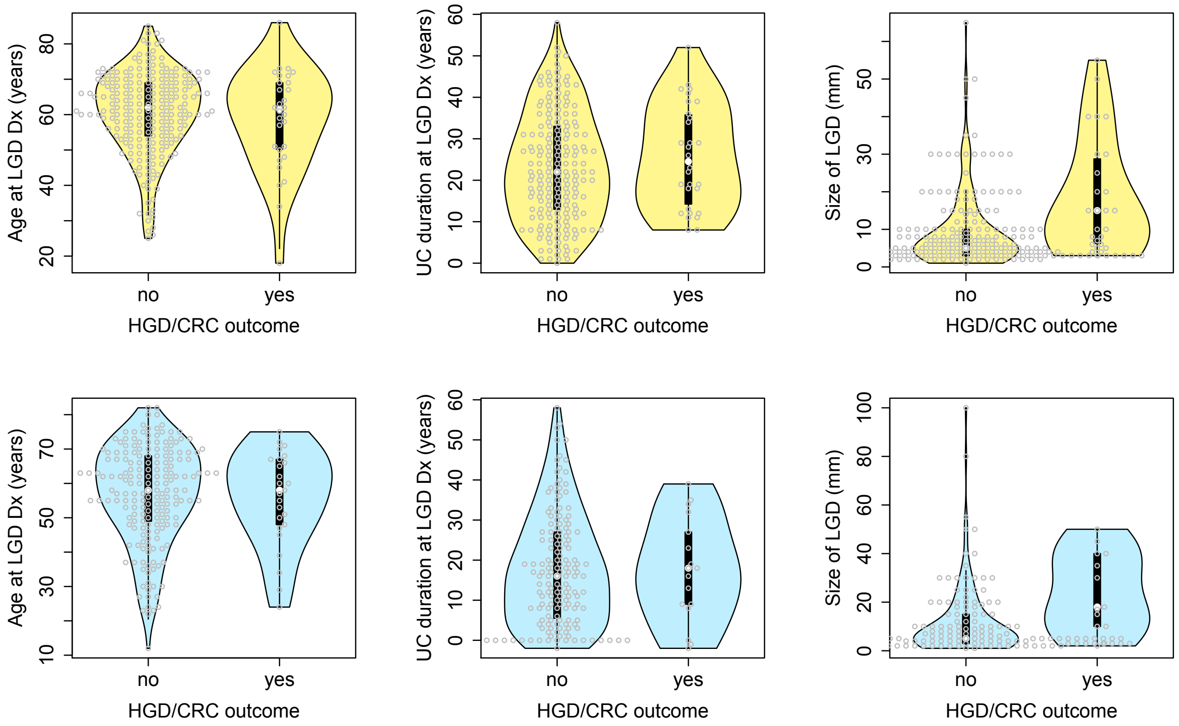


**Table S3: Multivariate model for progression to advanced neoplasia (AN) within the discovery set, stratified by pre vs post 2010 LGD diagnosis.**

Risk factors for low-grade dysplasia (LGD) progression to high-grade dysplasia or colorectal cancer (MULTIVARIATE Cox regression analysis). N=246 total patients included with data available, 29 progressed to AN. Score (logrank) overall p=9e-10 for model.

| Risk factor in final model | Hazard ratio (95% CI) | P value |
|---|---|---|
| **Visible index LGD size 10mm or more** | 2.7 (1.2 – 6.1) | 0.01288 |
| **Index LGD not endoscopically resected or incomplete resection** | 3.3 (1.4 – 7.5) | 0.00449 |
| **Multifocal LGD at time of index LGD diagnosis** | 2.9 (1.3 – 6.3) | 0.00734 |
| **Moderate or severe active histological inflammation in any colonic segment at time of or within previous 5 years of index LGD diagnosis** | 3.2 (1.5 – 6.8) | 0.00285 |

**Figure S2 : Baseline cumulative hazard functions estimated for stratified multivariate Cox proportional hazards analysis in Discovery set**. Solid blue and green lines show similar regression fits over first ten years of follow-up.

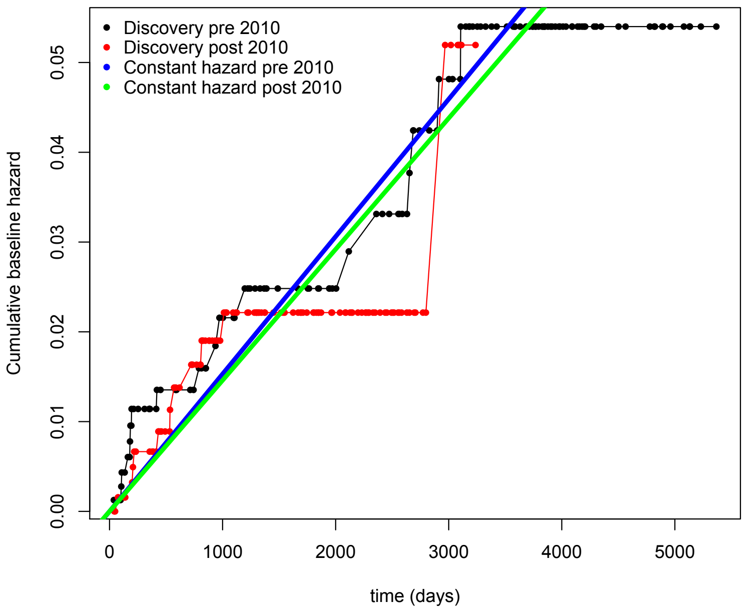


**Figure S3: Kaplan-Meier plots for probability of remaining free of high-grade dysplasia (HGD) or colorectal cancer (CRC) to assess risk stratification and predictive power of multivariate model.** (A) Discovery (n=246) and (B) validation (n=198) cohorts stratified by risk score (0-3+) defined by final multivariate model at index low-grade dysplasia (LGD) diagnosis to 14.7 years and 13 years of total available follow-up since index LGD in the discovery (A) and validation (B) cohorts, respectively.

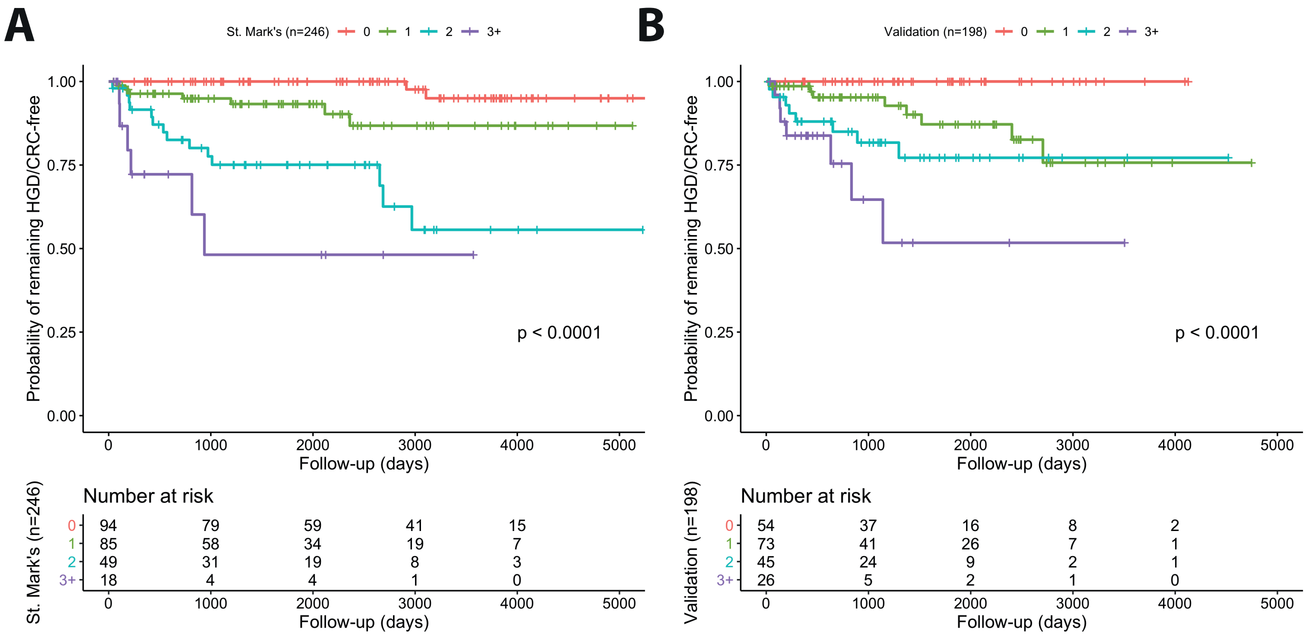

**Figure S4: Kaplan-Meier plots for probability of remaining free of high-grade dysplasia (HGD) or colorectal cancer (CRC) to assess risk stratification and predictive power of multivariate model in the recent data (2010 and later).** Modern cohort stratified by risk score (0-3+) defined by final multivariate model at index low-grade dysplasia diagnosis over total years of patient follow-up, patients included across both datasets.

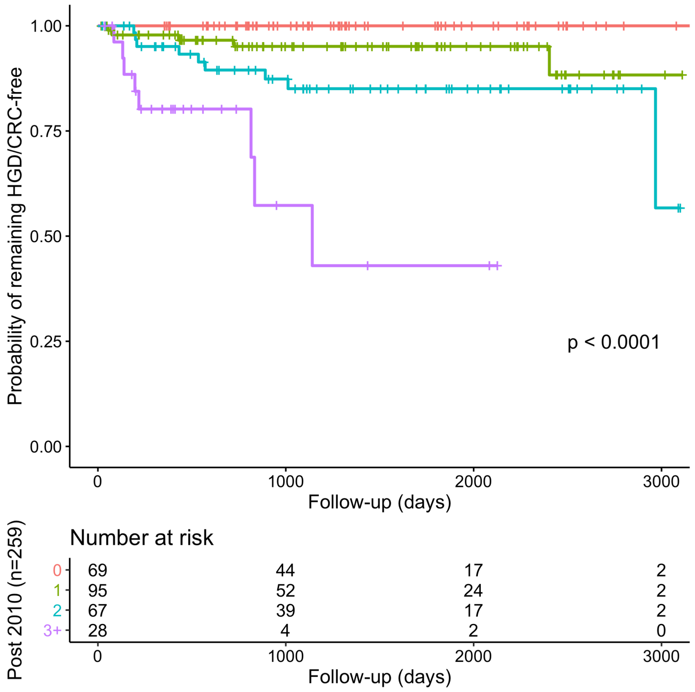


**Table S4: Predictive accuracy in highest and lowest risk groups in discovery and validation sets.** Positive predictive values (PPV) and negative predictive values (NPV) shown for each risk tier (i.e., for patient groups with scores 0, 1, 2, 3+ for lowest to highest predicted risk groups) in discovery and validation cohorts (rows) attained by designated time of follow-up (columns). Values derived from Kaplan-Meier estimation depicted in Figure S3.

| Group | At 6 months | At 1 year | At 3 years | At 5 years | At 10 years |
|---|---|---|---|---|---|
| **Discovery NPV** Risk score 0 | 100% | 100% | 100% | 100% | 95% |
| **Discovery NPV** Risk score 1 | 98% | 96% | 95% | 93% | 87% |
| **Discovery NPV** Risk score 2 | 96% | 92% | 75% | 75% | 56% |
| **Discovery NPV** Risk score 3+ | 87% | 72% | 48% | 48% | 48% |
| **Discovery PPV** Risk score 0 | 0% | 0% | 0% | 0% | 5% |
| **Discovery PPV** Risk score 1 | 2% | 4% | 5% | 7% | 13% |
| **Discovery PPV** Risk score 2 | 4% | 8% | 25% | 25% | 44% |
| **Discovery PPV** Risk score 3+ | 13% | 28% | 52% | 52% | 52% |
| **Validation NPV** Risk score 0 | 100% | 100% | 100% | 100% | 100% |
| **Validation NPV** Risk score 1 | 99% | 99% | 95% | 87% | 76% |
| **Validation NPV** Risk score 2 | 95% | 88% | 82% | 77% | 77% |

| | | | | | |
|---|---|---|---|---|---|
| **Validation NPV** Risk score 3+ | 88% | 84% | 65% | 52% | 52% |
| **Validation PPV** Risk score 0 | 0% | 0% | 0% | 0% | 0% |
| **Validation PPV** Risk score 1 | 1% | 1% | 5% | 13% | 24% |
| **Validation PPV** Risk score 2 | 5% | 12% | 18% | 23% | 23% |
| **Validation PPV** Risk score 3+ | 12% | 16% | 35% | 48% | 48% |

***UC-CaRE* risk prediction model development using discovery data**

Within the *UC-CaRE* web-tool we embedded a prognostic risk function based on the calibrated multivariate model that takes the values of the specified endoscopic features (risk factors) included in the risk score as input parameters for a patient with LGD and first stores them in the variable vector **x.** The estimated linear predictor coefficients $\beta = \log{(HR)}$ associated with each risk factor in the multivariate model were extracted and fixed from the output of the Cox proportional hazards analysis using the discovery set.

To calculate a patient's time-dependent risk of advanced neoplasia (AN), *UC-CaRE* calculates the probability $\Pr{(T_C < t)}$ where $T_C$ is the time of advanced neoplasia (AN) development, and $t$ the follow-up time since baseline LGD diagnosis. With $S(t)$ as the survival function for AN occurrence by time t, we have:

$$\Pr(T_c < t) = 1 - S(t|x) = 1 - \exp\left(-\hat{\Lambda}(t)\right)$$

$$\Rightarrow \text{Risk}(t) = 1 - \exp\left(-\exp(\beta \cdot x)\hat{\Lambda}_0(t)\right)$$

where $\widehat{\Lambda}_0(t)$ is the smoothed baseline cumulative hazard function obtained from the multivariate model fit to the discovery dataset. With the estimated covariance matrix and point estimates for the coefficients from Cox regression output (see Table S7 below), parametric bootstrapping was performed by sampling from a multivariate normal for beta coefficients, then computing the patient-specific *Risk(t)* defined above for each bootstrap iteration, and calculating confidence intervals for the risk predicted from quantiles of the probability estimates.

For the baseline hazard, we noted that the cumulative (non-parametric) baseline hazard estimated in the discovery set (n=246) was approximately linear, similarly in the validation set, implying a constant hazard rate of AN progression in LGD, particularly in the first 10 years following LGD diagnosis (Figure S5). Noting that the majority of total patient follow-up occurred by year 10 in our study, we used the discovery set to estimate the constant hazard rate from the origin at time 0 to the value of the baseline hazard at year 10 in yearly increments using linear regression. Thus, we assumed the following form for the baseline cumulative hazard function in our model, $\widehat{\Lambda}_0(t) = \lambda t$, where the constant hazard rate to progress to AN was estimated to be $\lambda$ = 0.0053 (95% CI 0.0047 – 0.0058) per year. We applied this smoothed baseline hazard as fixed in our prognostic *Risk(t)* function (Materials and Methods). As a potential alternative to the Cox model for use in *UC-CaRE*, we also explored fitting an exponential (equivalently, Poisson regression) model that results directly from the constant baseline hazard assumption (see below for model details and Table S5 for results).

**Figure S5: Baseline hazard functions derived with R *survival* package in discovery and validation sets.** Blue line shows linear regression for a smoothed constant hazard rate from 0 to 10 years of follow-up in discovery data.

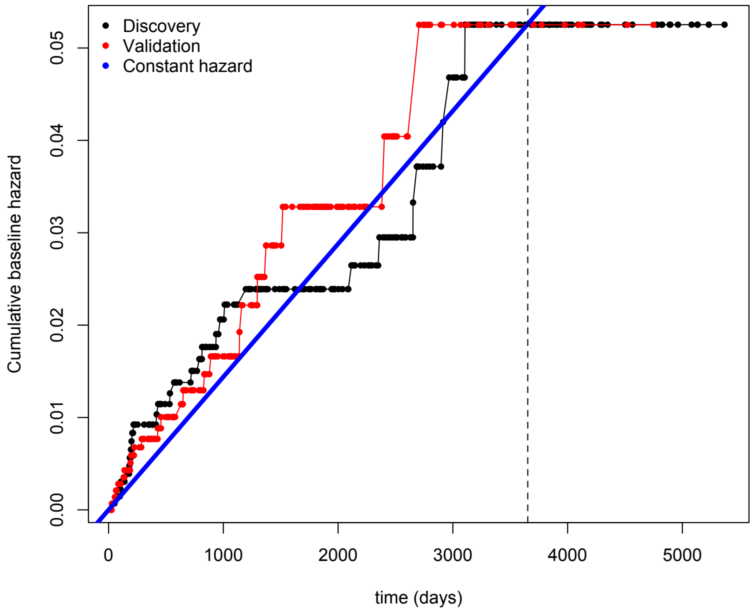


**Evaluation of *UC-CaRE* risk predictions for advanced neoplasia (AN) in validation data**

We can test the prognostic power of our model by comparing the observed (O) versus expected (E) numbers of AN progressors during follow-up in the validation set which requires computation of the cumulative hazard $\widehat{\Lambda}$(t) from our model (with HRs derived solely from the discovery set) in *UC-CaRE*. We considered both the total number across all patient risk groups and also the number predicted for each of the *k* risk groups defined by risk factor count separately. We compute the total cumulative hazard function as follows,

$$\hat{\Lambda}(t) = \sum_{i=1}^{k} \hat{\Lambda}_k(t) = \sum_{i=1}^{k} \sum_{i=1}^{N_k} \exp(\beta \cdot x_i) \hat{\Lambda}_0(t)$$

where $N_k$ is the total number of patients in risk score group *k* and $t_i$ is the patient person-days follow-up time to AN progression or censoring by time *t* for patient *i*. The above methods are based on cumulative AN progression-specific hazards and may be used to assess calibration both overall and in risk score subgroups of the validation data. We constructed a test assuming that O is generated from a Poisson distribution, whose rate would be E if the model was perfectly calibrated. Exact Poisson confidence intervals on O may be used to compute a confidence interval for O/E, the standardised incidence ratio[1].

To validate the tool's predictions, we used the prognostic function to predict risk in the validation patient data. Using all event data in the validation set, we computed the expected number Expected(t) = 1 + 5 + 7 + 10 = **23 AN cases** versus the observed number which was Observed(t) = 0 + 8 + 8 + 7 = **23 AN cases** from baseline until time of last follow-up exam (t= 13 years). Using an exact Poisson confidence interval for the observed (O=23, 95% CI = 14.6 - 34.5)[2], we estimated overall calibration in the large that can be expressed as a standardized incidence ratio, O/E = 1 (95% CI 0.63 - 1.5).

**Figure S6: Two methods of validation *UC-CaRE*** – 1) Predicted future risk of HGD/cancer (AN) in individual LGD patients (blue lines) since time of diagnosis based on multivariate model coefficients in *Risk(t)* function for each patient (see Materials and Methods). Green diamonds indicate observed (O) AN cases within each risk score group, blue shaded regions are interquartile confidence intervals for patient-specific predictions. Overall calibration is O/E = 1 (95% CI 0.63 - 1.5). 2) Positive predictive values (PPV) and negative predictive values (NPV) for model at certain follow-up years in validation set by Kaplan-Meier analysis (see Results and Figure S7 below).

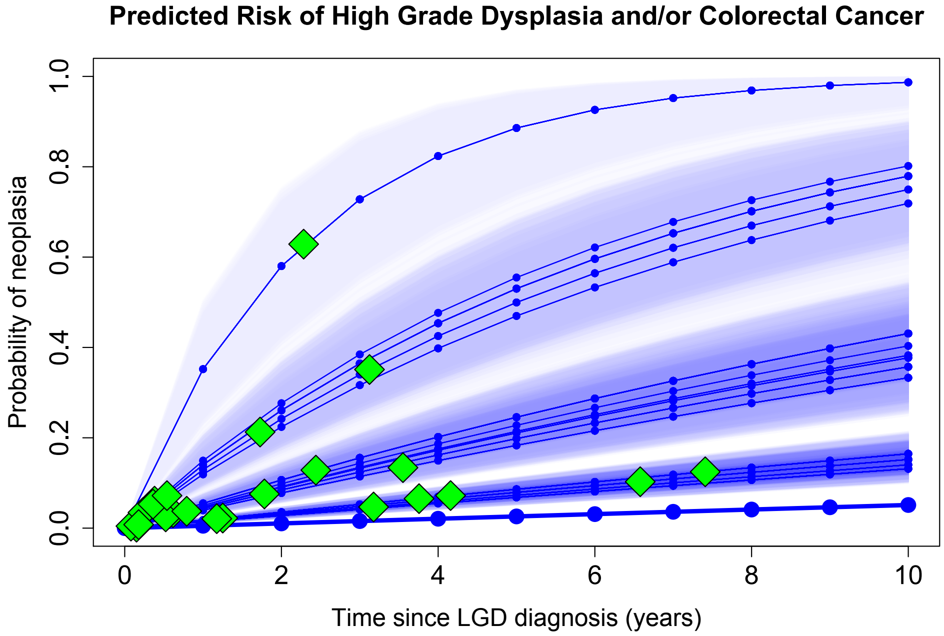


**Figure S7: Kaplan-Meier plots for probability of remaining free of high-grade dysplasia (HGD) or colorectal cancer (CRC) to assess risk stratification and predictive power of multivariate model.** Discovery (n=246) and validation (n=198) cohorts stratified by 5 risk score groups (0-4, top row) and 3 risk score groups (0, 1-2, 3+, bottom row) defined by final multivariate model at index low-grade dysplasia (LGD) diagnosis over total years of patient follow-up.

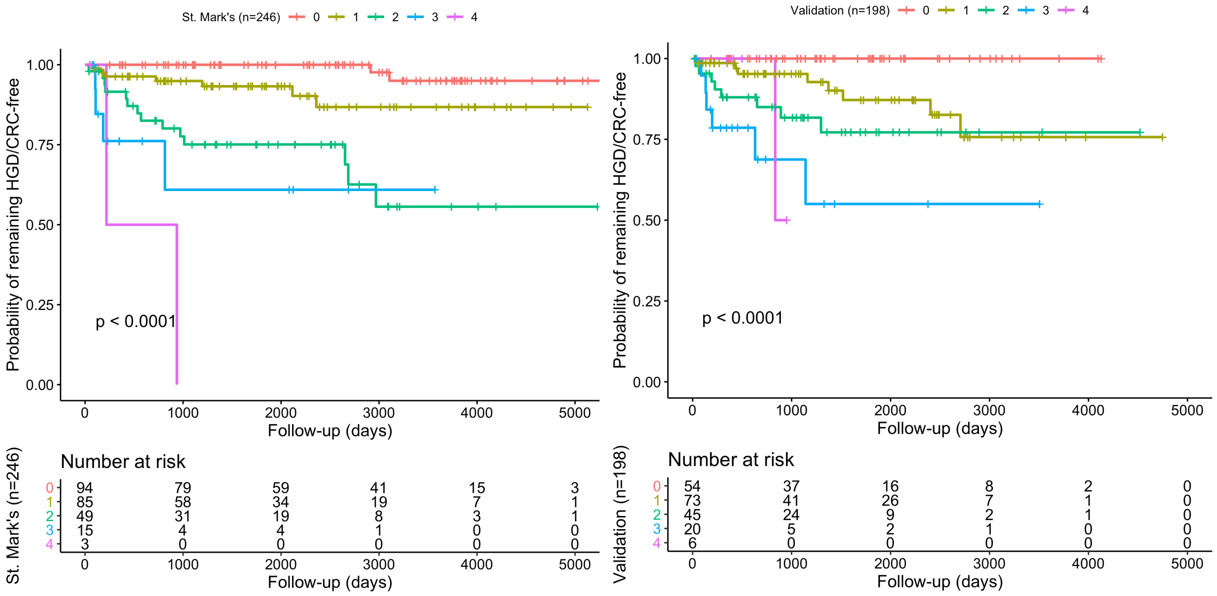

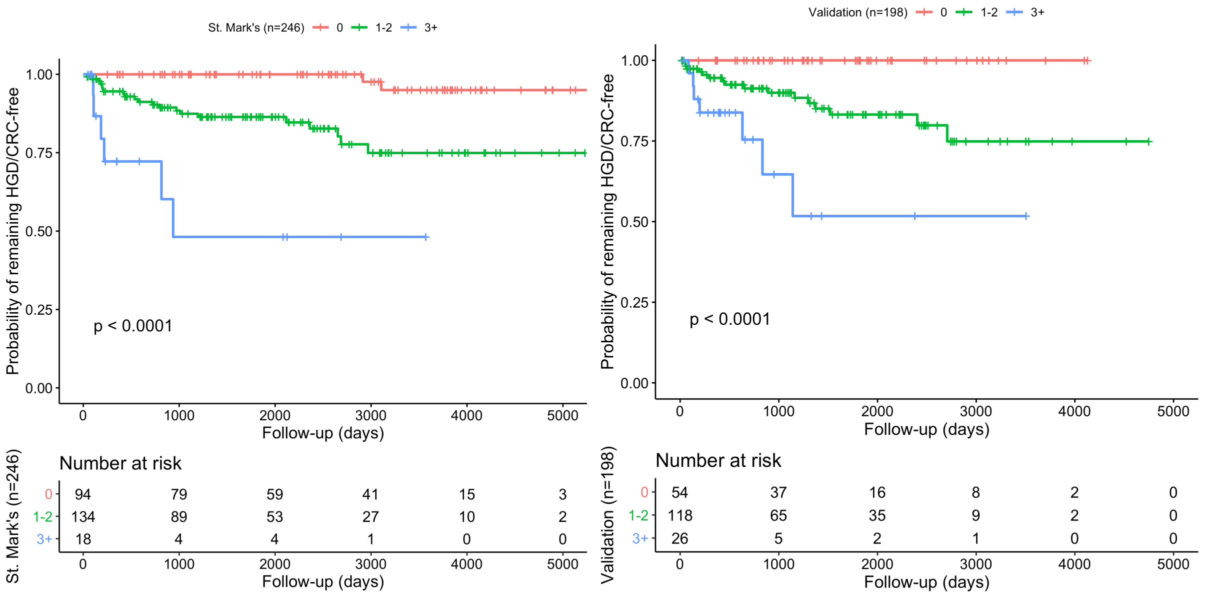


**Poisson regression model for constant hazard assumption**

With the assumption of a constant baseline hazard rate, we derived the expected number of advanced neoplasia cases as the following cumulative incidence function for *k* risk groups to use in model calibration,

$$\hat{\Lambda}_k(t) = \sum_{i}^{N_k} \exp(\beta \cdot x_i)\hat{\Lambda}_0(t)$$
$$= \exp(\beta \cdot x_1) \cdot \lambda \cdot t_1 + \exp(\beta \cdot x_2) \cdot \lambda \cdot t_2 + \ldots + \exp(\beta \cdot x_N) \cdot \lambda \cdot t_N$$

We note that the prognostic Risk function in the Main Text becomes an exponential model and thus an alternative method would be to fit a Poisson regression model to our discovery data and employ the output in the functional components of *UC-CaRE*. We summarize the results fitting such a model in Table S5. Although the baseline hazard was predicted to be similar ( exp(Intercept) ), we found that the hazard ratios (HRs) of this model for the selected risk factors are greater than those found in our main Results using a multivariate Cox proportional hazards (PH) model. During model calibration, the effects of this are seen in an over-estimate of the predicted number of cases in the validation set, as computed above, to be Expected(t) = 1 + 5 + 7 + 12 = **25 advanced neoplasia (AN) cases** versus the 23 observed. Thus, in the main Results, we chose to retain the original, more accurate model using a smoothed baseline hazard and HRs determined from the multivariate Cox PH model for use in the prognostic risk function employed in *UC-CaRE* for prospective patient assessment.

**Table S5. Poisson regression model for progression to advanced neoplasia within the discovery set**
Risk factors for LGD progression to HGD or CRC (MULTIVARIATE Poisson regression analysis)

| Variable in final model | Exp(coeff) (95% CI) | P value |
|---|---|---|
| **Baseline hazard rate [ exp(Intercept) per year]** | 0.005 (0.002 - 0.009) | <2e-16 |
| **Visible index LGD size 10mm or more** | 2.9 (1.3 – 6.3) | 0.00721 |
| **Index LGD not endoscopically resected or incomplete resection** | 3.9 (1.8 – 8.4) | 0.00042 |
| **Multifocal LGD at time of index LGD diagnosis** | 3.2 (1.5 – 7.1) | 0.00302 |
| **Moderate or severe active histological inflammation at time of or within previous 5 years of index LGD diagnosis** | 3.3 (1.6 – 7.1) | 0.00167 |

**Competing risks analysis**

In the Main Text, we note that the analyses performed using univariate and multivariate Cox proportional hazards (PH) models pertained to the cause-specific hazard (CSH) of progression to advanced neoplasia (AN) as an event separate to the event of colectomy, the latter of which served as a condition for censoring. In each of the patient datasets analysed, censoring due to colectomy occurred to 11% of patients during the study period (27/249 in discovery set and 24/211 in validation set). In practice, colectomy surgeries may be performed due to medically refractory symptomatic colitis, a cancer diagnosis or if there is a high risk of cancer in the future. The aim of our study is to determine which patients diagnosed with low-grade dysplasia (LGD) are considered to be at a considerable risk of progression to advanced neoplasia (AN) and thus a recommendation for colectomy in their case is justified. We acknowledge that censoring patients at colectomy before they have had time to progress to AN may affect estimates of overall absolute risk. Therefore, we performed additional analyses including competing risk of colectomy on study and summarize the results below.

First, we considered event-free survival method for Kaplan-Meier (KM) estimation, which focuses on the non-occurrence of events (event-free status, Figure S8A). In the KM plot produced using R package *cmprsk* for competing risks, the curve estimates the proportion of patients within each risk factor set (see Main Text) who have not experienced either of the endpoints since baseline LGD diagnosis (colectomy nor AN progression) in the discovery set. We noted that the curves are qualitatively similar to those using the same stratification for cause-specific hazard estimation, implying that the more high-risk patients designated by our risk factor criteria were also those more likely referred to colectomy. Furthermore, the cumulative incidence functions (CIFs) for both events of AN progression (event = 1) and colectomy (event = 2) are estimated to be very similar in the first years of follow-up in the presence of the competing risk event (Figure S8B created with function 'ggcompetingrisks' in R package *survminer*). This suggests patients received colectomy at a similar rate to advancing to neoplasia in our discovery set; patients who were at such high-risk for AN progression likely should have all been assigned the group who received colectomy during the study period if they were able.

Lastly, we graphically compared the CIFs and 1 - CSH KM curves in Figure S8C. The 1 - CSH KM (dotted lines) are nearly identical for the bottom 2 risk groups but overestimate the progression to AN in the top 2 risk groups in later follow-up years. Again, CIF curves (solid and dashed lines by event type AN progression and colectomy, respectively) describe the occurrence of each event type in the presence of the other, i.e., it describes the outcomes for each risk group in the data, and Gray tests were performed that found significant differences in risk of either event by risk score group ($p < 10^{-6}$ for both events)[3]. The 1 - CSH KM curves describe the progression risk for one type of event in the absence of the other, which is relevant for inference of the biological relationship between a certain event and given covariates. It overestimates the incidence because in the absence of the competing risk of a colectomy event, the event has more chance to occur. Overall, we found using competing risk formulation that the risks were similar for colectomy and AN progression in our discovery set, validating the hazard ratios used in our prognostic risk function in *UC-CaRE*.

**Table S6. Multivariate model for cause-specific progression to colectomy outcome within the discovery set**. Risk factors for occurrence of colectomy (MULTIVARIATE Cox regression analysis). N=246, including 27 who received colectomy as outcome for censoring.

| Risk factor in final model | Hazard ratio (95% CI) | P value |
|---|---|---|
| **Visible index LGD size 10mm or more** | 4.1 (1.8 – 9.2) | 0.0008 |
| **Index LGD not endoscopically resected or incomplete resection** | 5.8 (2.6 – 12.6) | 1.14e-05 |
| **Multifocal LGD at time of index LGD diagnosis** | 1.1 (0.5 – 2.5) | 0.8726 |
| **Moderate or severe active histological inflammation at time of or within previous 5 years of index LGD diagnosis** | 1.7 (0.8 - 3.8) | 0.1738 |

**Figure S8. Competing risk analysis of advanced neoplasia (AN) and colectomy within the discovery set.** (A) Event-free survival (combined colectomy and AN progression) derived from Kaplan-Meier estimation in discovery set (n=246) using same risk score groups from Main Text. (B) Cumulative incidence functions in the face of competing risks for event 1 (AN progression) and event 2 (colectomy) were found to be similar in this cohort. (C) Event outcomes in the data (CIFs) and model estimated survival (1 – CSH) by risk factor groups.

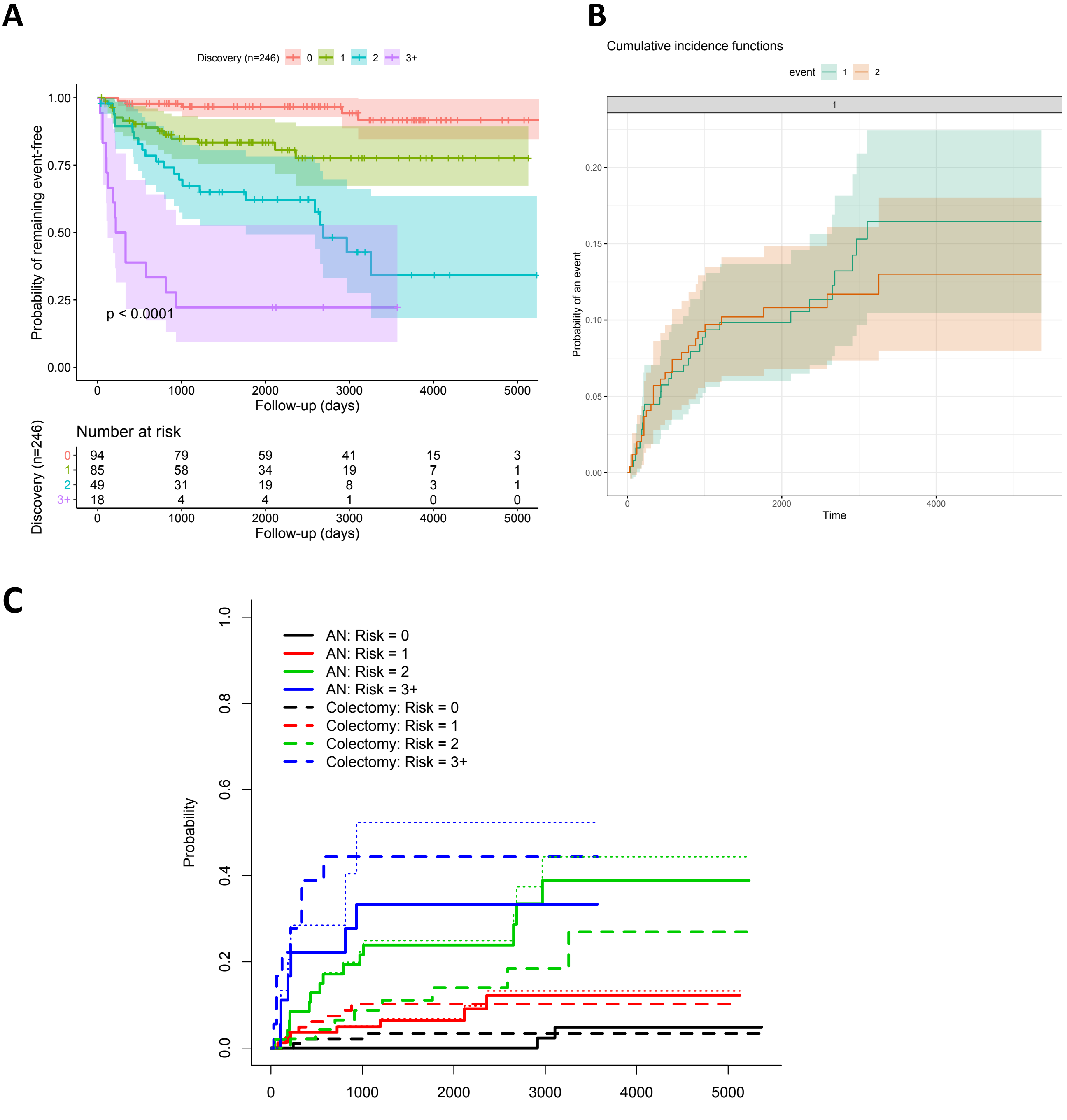


**Table S7. Variance-covariance matrix for final multivariate model for cause-specific progression to advanced neoplasia (AN) outcome derived from the Discovery Set**. Risk factors for occurrence of colectomy (MULTIVARIATE Cox regression analysis; SE = standard error for adjusted coefficient in model)

| Risk factor in final model | 1 | 2 | 3 | 4 |
|---|---|---|---|---|
| **1. Visible index LGD size 10mm or more (SE: 0.4013)** | 0.161051423 | -0. 005849893 | -0.046865593 | -0. 001038021 |
| **2. Index LGD not endoscopically resected or incomplete resection (SE: 0.3959)** | -0.005849893 | 0.156726196 | -0.005749705 | -0.026911947 |
| **3. Multifocal LGD at time of index LGD diagnosis (SE: 0.3945)** | -0.046865593 | -0.005749705 | 0.155634218 | 0.020300342 |
| **4. Moderate or severe active histological inflammation at time of or within previous 5 years of index LGD diagnosis (SE: 0.3856)** | -0.001038021 | -0.026911947 | 0.020300342 | 0.148660355 |

**Figure S9: : Kaplan-Meier plot for probability of remaining HGD/CRC free to assess differences in risk between patients with Incomplete resection such that highest risk lesion was invisible (code = 2; i.e., no unresectable non-polypoid lesions) versus Incomplete resection such that highest risk lesion was not invisible (code = 1)** in the validation (n=198) cohort.

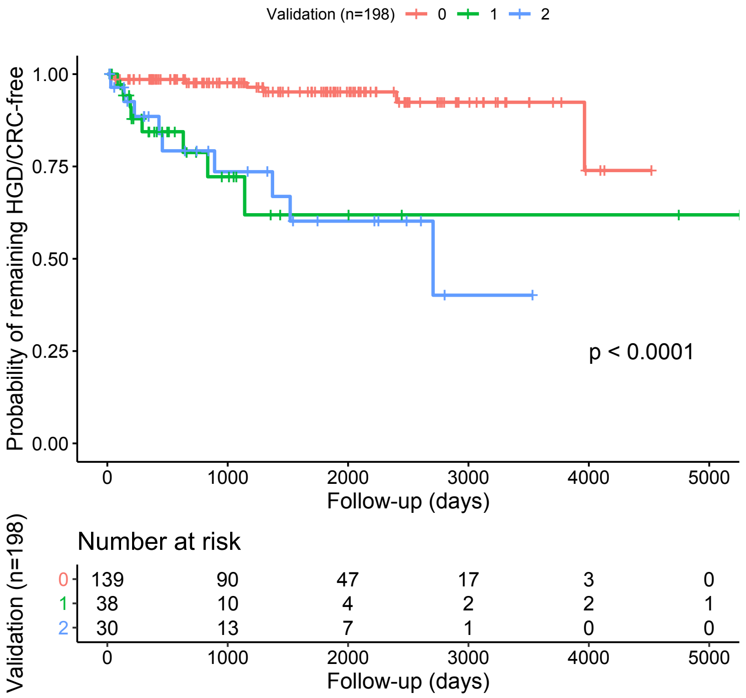


##### Patient and Public Involvement

Patients were not directly involved in the retrospective design and conduct of this study but previous qualitative work involving patient questionnaires and interviews revealed aspects that we aimed to include in the construction of our visual web-tool *UC-CaRE*. To summarize, patients described both a lack of quantitative risk communication and a lack of personalised discussion of treatment options as barriers to patient decision-making.[4] Thus we included calculations of such personalised, quantitative risks (percentages by follow-up year) in easy to interpret visual diagrams (Paling charts) as part of this study. We plan to disseminate our conclusions to patients by implementing UC-CaRE for use in shared treatment decision-making and obtaining feedback/critique on their experiences for future improvements.
